## Supplementary Appendix for "The cost and cost-effectiveness of novel tuberculosis vaccines in low- and middle-income countries: a modelling study"

### S1. Epidemiological methods

The subsequent pages provide relevant details and methods regarding the underlying epidemiological model and vaccine delivery scenarios from Clark et al. [1].

#### S1.1. Tuberculosis natural history structure

The core natural history model is specified in Figure S1.1. Those with no previous exposure or infection with *Mtb* [Uninfected-Naive ($U_N$)] could become infected at rate $\lambda_j$ and progress to an Infection-Fast ($I_F$) class following initial infection. From Infection-Fast, three possible pathways were possible: (i) Fast progression to Subclinical Disease ($D_S$), where individuals are infectious with a reduced infectiousness compared to clinical tuberculosis, but display no symptoms of tuberculosis disease [2]; (ii) self-clearance to Uninfected-Cleared ($U_C$), where individuals are no longer infected with Mtb and therefore are not at risk of progression to tuberculosis disease without reinfection [3]; or (iii) continue to remain latently infected with a risk of reactivation and progression to disease, albeit at a lower rate than Infection-Fast, by transitioning to the Infection-Slow ($I_S$) class. Those in the Infection-Slow class could self-clear to the Uninfected-Cleared class, be reinfected and return to the Infection-Fast class or reactivate their infection and progress to Subclinical Disease.

Once in the Subclinical Disease class, individuals could naturally cure (without treatment) to the Resolved (R) class, or progress to Clinical Disease ($D_C$), where individuals are infectious and display symptoms of tuberculosis disease. Treatment initiation from Clinical Disease to On-Treatment (T) began in 1960 and increased following a sigmoid curve to 2019, with average treatment duration assumed to be six months [4, 5]. Treatment completions transitioned to the Resolved class and treatment non-completions returned to Clinical Disease. Deaths occurring on-treatment and in clinical disease counted toward the total number of tuberculosis deaths during the year. Those with clinical disease could also naturally cure to the resolved class. Individuals in the Resolved class could be reinfected or relapse to Subclinical. We assumed that the infection and resolved classes are partially protected against reinfection [6, 7]. In those who have self-cleared, we assumed the level of protection against reinfection is half of the protection against reinfection for the infection and resolved classes. Age was modelled in single years from ages 0 to 79 and aggregated into two categories for ages 80 to 89, and ages 90 to 99. Births and ageing occurred at the beginning of each year.

#### S1.2. HIV and ART structure description

To account for the influences of human immunodeficiency virus (HIV) and antiretroviral therapy (ART) on the risk of infection with *Mtb* and progression to tuberculosis disease [6, 8], we have implemented an HIV structure (shown in Figure S2.1) composed of 3 compartments: HIV uninfected [HIV0], people living with HIV (PLHIV) not on ART [HIV1], and PLHIV on ART [ART]. HIV uninfected individuals were diagnosed with HIV and moved from the HIV0 compartment to the HIV1 compartment with rate $\lambda_H$. Within the HIV1 compartment, there is a higher risk of tuberculosis progression and an increased tuberculosis mortality rate compared to the HIV0 compartment. PLHIV are initiated on treatment with ART from HIV1 following a sigmoid trend. The increases in tuberculosis mortality rate and tuberculosis progression are reduced while in ART compared to HIV1, but still higher than in HIV0. ART also reduces the HIV mortality rate.

The separate stratum was included to dynamically model the tuberculosis-HIV co-epidemic if the proportion of tuberculosis cases among people living with HIV (PLHIV) was greater than or equal to 15%, and if the HIV prevalence in the country was greater than 1%. Countries incorporating the additional HIV structure are listed in Table S1.1.

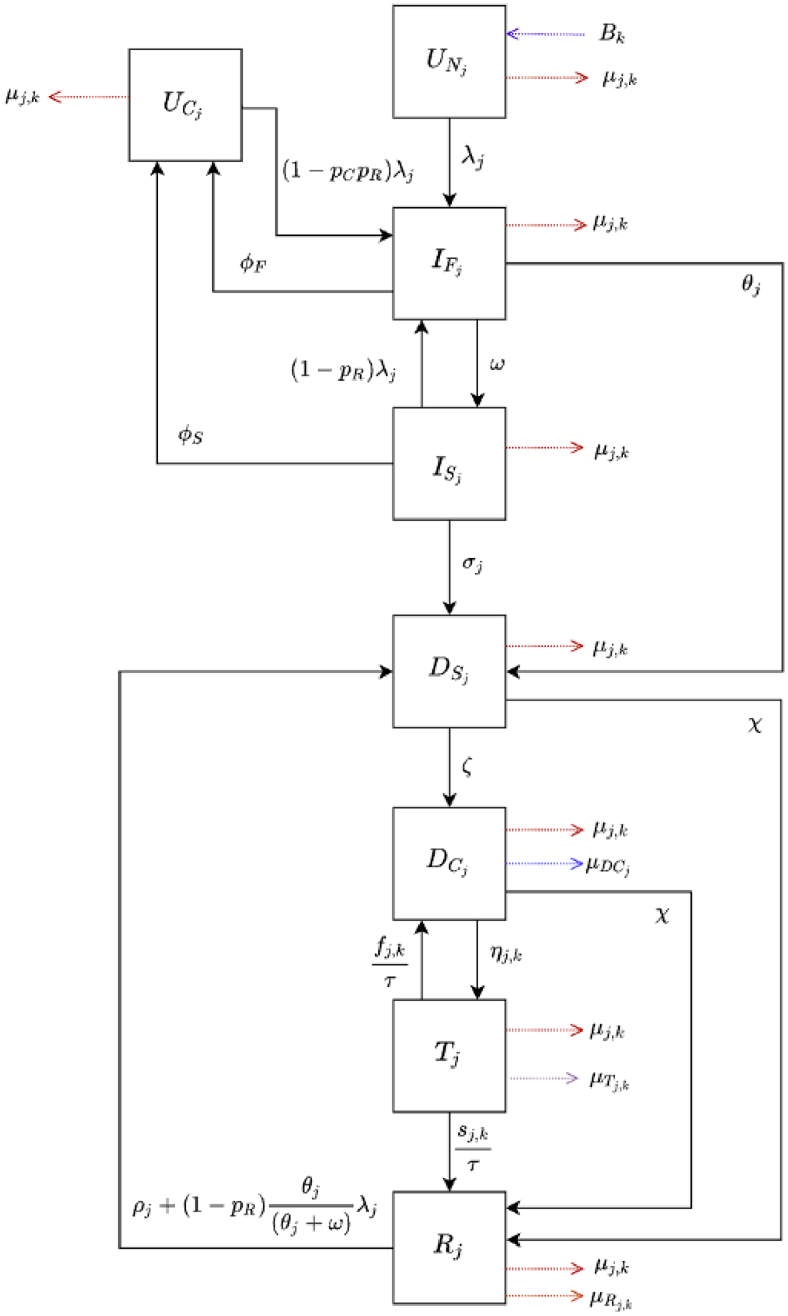


**Figure S1.1. Tuberculosis natural history model**

*Abbreviations: $D_C$ = Clinical Disease; $D_S$ = Subclinical Disease; $I_F$ = Infection-Fast; $I_S$ = Infection-Slow; R = Resolved; T = On-Treatment; $U_C$ = Uninfected-Cleared; $U_N$ = Uninfected-Naïve.*

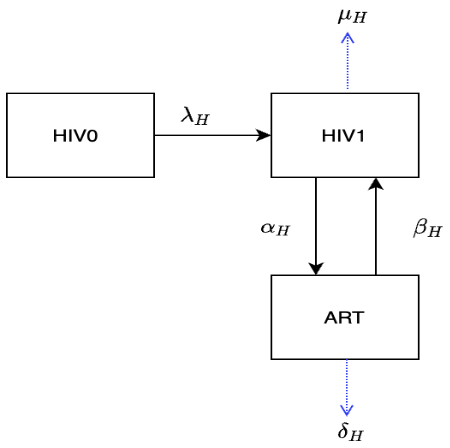


**Figure S2.1. HIV and ART structure.**

*Abbreviations: ART = People living with HIV on ART; HIV0 = HIV uninfected; HIV1= People living with HIV not on ART.*

**Table S1.1. Countries incorporating the HIV structure with their corresponding HIV prevalence and proportion of tuberculosis cases among PLHIV.**

| Country | HIV Prevalence (%) | Proportion of tuberculosis cases among PLHIV (%) |
|---|---|---|
| Botswana | 16.5 | 48.6 |
| Central African Republic | 2.1 | 25.4 |
| Côte d'Ivoire | 1.7 | 17.5 |
| Cameroon | 2.0 | 26.8 |
| Gabon | 2.3 | 32.8 |
| Ghana | 1.1 | 20.8 |
| The Gambia | 1.2 | 17.7 |
| Guinea-Bissau | 2.1 | 31.3 |
| Equatorial Guinea | 4.8 | 26.5 |
| Guyana | 1.1 | 19.0 |
| Kenya | 2.9 | 26.2 |
| Lesotho | 16.0 | 61.6 |
| Mozambique | 7.2 | 33.8 |
| Malawi | 5.9 | 46.6 |
| Namibia | 8.4 | 32.5 |
| Rwanda | 1.8 | 21.1 |
| Eswatini | 17.4 | 60.1 |
| Togo | 1.5 | 16.2 |
| Tanzania | 2.9 | 23.6 |
| Uganda | 3.4 | 39.0 |
| South Africa | 12.8 | 58.0 |
| Zambia | 6.7 | 46.2 |
| Zimbabwe | 9.6 | 59.8 |

### S1.3. Calibration methodology

The model was fitted to epidemiologic calibration targets using history matching with emulation, implemented using the hmer R package [9, 10]. If countries were unable to find at least 1000 fully fitted parameter sets using this method, they were subsequently assessed using an Approximate Bayesian Computation using Markov Chain Monte Carlo method (ABC-MCMC). ABC-MCMC was conducted using the easyABC package in R, modified by Sebastian Funk, Gwenan Knight, and the Tuberculosis Modelling group at LSHTM for adaptive sampling and to accept seeded parameter values [9, 11]. We used parameter sets with the maximum number of targets fitted using history matching with emulation as a starting seed, with the ABC-MCMC algorithm continuously adapting using the last 1000 points and the noise factor set to 0.0001.

Analysis was performed on 105 countries from the 135 total low- and middle-income countries identified based on 2019 World Bank Income groups. There were 20 countries excluded from the initial calibration attempt due to missing crucial data required to attempt calibration, and 10 countries which were unable to be calibrated (could not find a parameter set that matched all targets using both history matching with emulation as well as ABC-MCMC). Reasons for exclusion from the final list of calibrated countries are provided in Table S2.1.

**Table S2.1. Reasons for exclusion from the final list of calibrated countries.**

| Country | Reason for Exclusion |
|---|---|
| Algeria | Did not calibrate |
| American Samoa | Missing multiple critical epidemiological data for calibration, no contact matrices available |
| Belize | No case notification or incidence data for children |
| Bosnia and Herzegovina | Did not calibrate |
| Cabo Verde | Did not calibrate |
| Comoros | No case notification data |
| Democratic Republic of the Congo | No case notification data by age |
| Republic of the Congo | No population estimates |
| Democratic People's Republic of Korea | Missing multiple critical epidemiological data for calibration |
| Djibouti | No case notification data by age |
| Dominica | Missing multiple critical epidemiological data for calibration, no contact matrices available |
| Guinea-Bissau | Did not calibrate |
| Guyana | Did not calibrate |
| Federated States of Micronesia | Missing multiple critical epidemiological data for calibration, no contact matrices available |
| Grenada | Missing multiple critical epidemiological data for calibration, no contact matrices available |
| Haiti | Missing 2020 contact matrix |
| Jamaica | Did not calibrate |
| Kiribati | Missing 2020 contact matrix |
| Kosovo | Missing multiple critical epidemiological data for calibration |
| Lebanon | Missing 2020 contact matrix |

| | |
|---|---|
| Marshall Islands | Missing multiple critical epidemiological data for calibration, no contact matrices available |
| North Macedonia | Did not calibrate |
| Samoa | No case notification or incidence data for children |
| Somalia | No contact matrices available |
| St. Lucia | No case notification or incidence data for children |
| St. Vincent and the Grenadines | Did not calibrate |
| Tonga | Did not calibrate |
| Turkmenistan | Did not calibrate |
| Tuvalu | No contact matrices available |
| West Bank and Gaza | Missing multiple critical epidemiological data for calibration |

### S1.4. Vaccine profile

The vaccine profile for an adult/adolescent vaccine and infant vaccine were based on the WHO Preferred Product Characteristics for New Tuberculosis vaccines [12], and are outlined in Table S3.1 below.

**Table S3.1. WHO Preferred Product Characteristics for New Tuberculosis Vaccines.**

| Vaccine | Host infection status at time of vaccination required for efficacy | Effect type | Vaccine efficacy | Duration of protection |
|---|---|---|---|---|
| Adolescent / Adult | Pre- and post-infection | Prevention of disease | 50% | Lifelong |
| | | | | 10 years |
| Infant | Pre-infection | Prevention of disease | 80% | Lifelong |
| | | | | 10 years |

Vaccine efficacy was assumed to be the same in both PLHIV and HIV-naïve recipients in countries incorporating the HIV structure, and in both younger age groups and older adults. The vaccine was assumed to have the same impact on preventing drug-susceptible and drug-resistant tuberculosis as specified in the WHO PPCs [12]. As we were modelling a prevention of disease vaccine, there was no direct impact on *Mtb* transmission or the force of infection.

We assumed duration of protection was 10 years on average, in addition to a sensitivity analysis with lifelong duration of protection. The shape of waning immunity was modelled as an exponential distribution, based on similar shapes for waning vaccine immunity of BCG [13] and other vaccines [14, 15].

#### S1.5. Vaccine delivery scenarios

The infant vaccine was implemented in two scenarios, and, separately, the adolescent/adult vaccine was implemented in three scenarios. The *Basecase* and *Accelerated Scale-up* scenarios included routine single-dose neonatal vaccination for the infant vaccine (85% coverage), and routine single-dose vaccination of 9-year-olds (80% coverage) with a one-time vaccination campaign for ages ten and older (70% coverage) for the adolescent/adult vaccine. The *Routine Only* scenario (adolescent/adult vaccine only) was introduced through routine 9-year-old vaccination only (i.e., no campaign). Specifics of the infant and adolescent/adult vaccine scenarios are provided in Table S4.1.

**Table S4.1. Vaccine scenarios for the infant and adolescent/adult vaccines**.

| **Characteristics** | **Infant Vaccine Scenarios** | | **Adolescent/Adult Vaccine Scenarios** | | |
|---|---|---|---|---|---|
| | ***Basecase*** | ***Accelerated Scale-up*** | ***Basecase*** | ***Accelerated Scale-up*** | ***Routine Only*** |
| **Ages Targeted** | *Neonatal:* Routine | *Neonatal:* Routine | *Age 9:* Routine<br><br>*Ages 10+:* One-time vaccination campaign over 5 years | *Age 9:* Routine<br><br>*Ages 10+:* One-time vaccination campaign in 2025 | *Age 9:* Routine |
| **Introduction Year** | Country-specific | 2025 | Country-specific | 2025 | Country-specific |
| **Vaccine Rollout Trend** | 5-year linear scale-up to coverage | Instant scale-up to coverage | 5-year linear scale-up to coverage | Instant scale-up to coverage | 5-year linear scale-up to coverage |
| **Target Coverage *(Low/Med/High)*** | 75% / 85% / 95% | | Age 9: 70% / 80% / 90%<br>Ages 10+: 50% / 70% / 90% | | |

### S1.6. Country-specific introduction years

In the *Basecase* and *Routine Only* scenarios, vaccines were introduced in country-specific introduction years between 2028 and 2047. The year 2028 was selected as the earliest country-specific introduction year to align with the anticipated completion and availability of results from TB vaccine candidate trials based on expert consultation and analysis. Country-specific introduction years were calculated for all 135 LMICs based on the 2019 World Bank Income groups. To calculate the specific year of introduction, countries were divided into two general categories: those procuring with support from Gavi, the Vaccine Alliance, and those self-procuring. Determination of country status was based on eligibility information posted on Gavi's website [16]. Countries transitioning from Gavi support are able to benefit from Gavi pricing and incremental financing for a period of 5–10 years. For countries that have already initiated the period of transition by 2019, this window will have largely ended by the time of tuberculosis vaccine availability through Gavi. As such, these countries were categorised as self-procuring countries. Countries that have not yet commenced transition, including India and Nigeria, were categorised as Gavi supported countries, given the long grace period post-commencement of transition. For more information, please see Gavi, https://www.gavi.org/types-support/sustainability/transition (retrieved December 1, 2020).

Through a consultative process with experts from WHO, Gavi, PATH, PDVAC, CHAI, and industry partners, factors influencing likelihood of being an early or late adopter were identified for both Gavi and self-procuring countries. Identified factors include disease burden, immunization capacity, and early adopter status. Country-specific registration timelines and commercial prioritization were also deemed important determinants of introduction timing for self-procuring countries.

*Additional factors for Gavi countries:* For countries procuring through Gavi, timelines for introduction are also influenced by Gavi processes. Prior to offering a new vaccine, Gavi requires that products be licensed, included in Gavi's Vaccine Investment Strategy, reviewed by SAGE, recommended in a WHO position paper, WHO prequalified, and approved for procurement by Gavi (Table S5.1). In addition, time for country application processing, contracting, and delivery must be factored. Through consultations, it was determined that a baseline time of roughly two years post licensure would be needed for Gavi processes prior to first country introduction, assuming several steps advance in parallel.

**Table S5.1. Timelines for Gavi processes post licensure.**

| | **Cumulative additional time (years)** | | |
|---|---|---|---|
| **Activities post licensure** | **Low End** | **High End** | **Average** |
| WHO PQ | 0.25 | 1.00 | 0.63 |
| SAGE Policy Review & WHO Position Paper | 0.25 | 0.50 | 0.38 |
| Gavi Decision | 0.25 | 0.50 | 0.38 |
| National review & Country applications | 0.25 | 0.75 | 0.50 |
| Contracting & delivery | 0.25 | 0.50 | 0.38 |
| **Years** | **1.25** | **3.25** | **2.25** |

*Weight of criteria, indicators, and scoring*: Differential weight was assigned to criteria based on their relative impact on the order of country adoption. This weight varied for self-procuring and Gavi countries (Table S6.1).

**Table S6.1. Weight of criteria influencing order of country adoption.**

| Criteria | Self-procuring countries | Gavi countries |
|---|---|---|
| Disease burden | 30% | 45% |
| Immunization capacity | 15% | 30% |
| Early adopter/leader | 15% | 25% |
| Lack of regulatory barriers | 15% | NA |
| Commercial prioritization | 25% | NA |

The following indicators were used to measure each of the variables identified in Table S7.1.

**Table S7.1. Indicators of criteria influencing order of country adoption.**

| Criteria | Indicator |
|---|---|
| **Disease burden** | Tuberculosis incidence |
| **Immunization capacity** | Proportion receiving 3 doses of DPT3 among infants 1 years of age (The percent of infants receiving 3 doses DPT3 is commonly used as a proxy for assessing immunization infrastructure) |
| **Lack of regulatory barriers** | Signatories to WHO PQ or SRA collaborative registration scheme<br>Lack of requirements for additional local clinical trial data |
| **Early adopter/leader** | Time to policy adoption of universal Xpert MTB/RIF screening for presumed tuberculosis cases<br>Time to adoption of HPV |
| **Commercial prioritization** | |
| *Ability to finance vaccines* | GDP per capita |
| *Political will to address tuberculosis* | Spending per tuberculosis case |
| *Market potential* | Population |

To standardize across these varied metrics, a point value ranging from 1–5 per criteria was assigned, with a score of 1 correlating with an earlier adopter and score of 5 correlating with a later adopter.

*Continuous variable*s such as disease burden or population were divided into quintiles. Those in the highest quintile were assigned a score of 1, those in the second highest quintile received a score of 2, and so forth. *Categorical variables* such as registration or early adopter status were scored based on whether countries met fixed criteria. For instance, countries that are signatories of WHO PQ or SRA collaborative registration schemes were assigned a score of 1. Those that are not signatories and have requirements for additional clinical trial data in local populations received a score of 5.

Scores were then weighted as reflected in Table S6.1 and aggregated into a composite score to determine countries' relative position in the queue of introductions.

Assumptions for the pace of introduction—i.e., how many countries per year would introduce the product and what the scale up curve might look like— was informed with data from pneumococcal vaccine (PCV) scale-up [17]. The percent of countries adopting each year (year 1 to year 12) for PCV was calculated. These annual percentages were then applied to tuberculosis vaccine scale up (based on a total n=135 countries: 78 self-procuring countries and 57 Gavi countries). The first year of tuberculosis vaccine scale up was estimated to be 2028, with Gavi countries following a similar scale up trajectory but delayed by two years due to required Gavi lead time for processing new vaccines (Table S5.1). Because PCV data is only available for 12 years, data was extrapolated for years 13 to 20 of tuberculosis vaccine roll out at a steady state. Country introduction timelines were adjusted—where applicable—to group countries with the same composite score in the same year of adoption. The cumulative number of countries introducing the vaccine by year is shown in Figure S3.1.

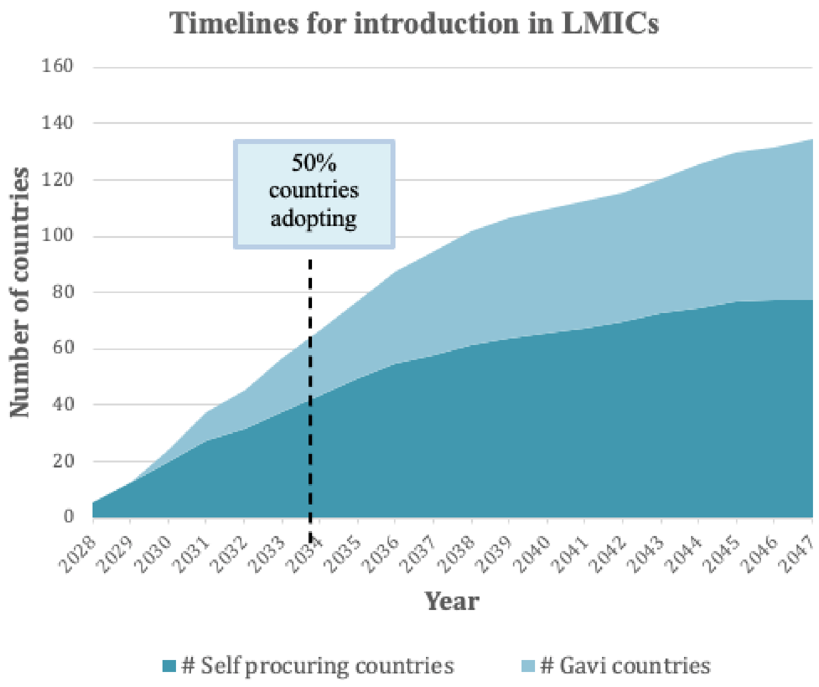


**Figure S3.1. Assumed cumulative number of countries introducing a novel vaccine per year.**

### S1.7. Vaccine coverage targets.

For each vaccine implementation scenario, low, medium, and high coverage targets for 5 years post-introduction were evaluated. The medium coverage target for the routine infant vaccination was 85%, based on the 2019 DTP3 (diphtheria, tetanus toxoid, and pertussis) average coverage level according to the WHO and UNICEF estimates of national immunisation coverage, with 10% uncertainty (low coverage = 75%, high coverage = 95%) [17]. Routine adolescent vaccination assumed a medium coverage target of 80% aligning with HPV coverage in South Africa combined with aggregated secondary school enrolment in China and India as assumed in Harris 2020 [18], also with 10% uncertainty targets (low coverage = 70%, high coverage = 90%). The medium coverage target for the adolescent/adult campaign was 70% aligning with the lower bound of the MenAfriVac campaigns in sub-Saharan Africa as assumed in Harris 2020 [18], with a wider uncertainty of 20% (low coverage = 50%, high coverage = 90%).

In the *Accelerated Scale-up* implementation, the 5-year coverage targets are achieved instantly in year 1, while in the *Basecase* and *Routine Only* implementations, the scale-up to coverage occurs linearly over 5 years.

### S2. Analyzed low- and middle-income country list.

| Country | WHO Region | Income level[a] | Gavi status | High-TB burden[b] | High-TB/HIV burden[b] | High-MDR/RR-TB burden[b] | Base-case vaccine introduction year |
|---|---|---|---|---|---|---|---|
| Afghanistan | EMR | LIC | Gavi | No | No | No | 2031 |
| Angola | AFR | LMIC | Gavi | Yes | No | Yes | 2032 |
| Albania | EUR | UMIC | Non-Gavi | No | No | No | 2035 |
| Argentina | AMR | UMIC | Non-Gavi | No | No | No | 2031 |
| Armenia | EUR | UMIC | Gavi | No | No | No | 2033 |
| Azerbaijan | EUR | UMIC | Gavi | No | No | Yes | 2028 |
| Burundi | AFR | LIC | Gavi | No | No | No | 2044 |
| Benin | AFR | LMIC | Gavi | No | No | No | 2037 |
| Burkina Faso | AFR | LIC | Gavi | No | No | No | 2039 |
| Bangladesh | SEAR | LMIC | Gavi | Yes | No | Yes | 2035 |
| Bulgaria | EUR | UMIC | Non-Gavi | No | No | No | 2029 |
| Belarus | EUR | UMIC | Non-Gavi | No | No | Yes | 2028 |
| Bolivia | AMR | LMIC | Gavi | No | No | No | 2037 |
| Brazil | AMR | UMIC | Non-Gavi | Yes | Yes | No | 2030 |
| Bhutan | SEAR | LMIC | Gavi | No | No | No | 2034 |
| Botswana | AFR | UMIC | Non-Gavi | No | Yes | No | 2028 |
| Central African Republic | AFR | LIC | Gavi | Yes | Yes | No | 2033 |
| China | WPR | UMIC | Non-Gavi | Yes | No | Yes | 2029 |
| Côte d'Ivoire | AFR | LMIC | Gavi | No | No | No | 2034 |
| Cameroon | AFR | LMIC | Gavi | No | Yes | No | 2031 |
| Colombia | AMR | UMIC | Non-Gavi | No | No | No | 2030 |
| Costa Rica | AMR | UMIC | Non-Gavi | No | No | No | 2033 |
| Cuba | AMR | UMIC | Gavi | No | No | No | 2035 |
| Dominican Republic | AMR | UMIC | Non-Gavi | No | No | No | 2031 |
| Ecuador | AMR | UMIC | Non-Gavi | No | No | No | 2033 |
| Egypt | EMR | LMIC | Non-Gavi | No | No | No | 2033 |
| Eritrea | AFR | LIC | Gavi | No | No | No | 2047 |
| Ethiopia | AFR | LIC | Gavi | Yes | Yes | No | 2030 |

| | | | | | | | |
|---|---|---|---|---|---|---|---|
| Fiji | WPR | UMIC | Non-Gavi | No | No | No | 2031 |
| Gabon | AFR | UMIC | Non-Gavi | Yes | Yes | No | 2038 |
| Georgia | EUR | UMIC | Gavi | No | No | No | 2029 |
| Ghana | AFR | LMIC | Gavi | No | No | No | 2040 |
| Guinea | AFR | LIC | Gavi | No | Yes | No | 2033 |
| Gambia | AFR | LIC | Gavi | No | No | No | 2039 |
| Equatorial Guinea | AFR | UMIC | Non-Gavi | No | No | No | 2042 |
| Guatemala | AMR | UMIC | Non-Gavi | No | No | No | 2036 |
| Honduras | AMR | LMIC | Gavi | No | No | No | 2037 |
| Indonesia | SEAR | UMIC | Gavi | Yes | Yes | Yes | 2034 |
| India | SEAR | LMIC | Gavi | Yes | Yes | Yes | 2033 |
| Iran | EMR | UMIC | Non-Gavi | No | No | No | 2031 |
| Iraq | EMR | UMIC | Non-Gavi | No | No | No | 2033 |
| Jordan | EMR | UMIC | Non-Gavi | No | No | No | 2037 |
| Kazakhstan | EUR | UMIC | Non-Gavi | No | No | Yes | 2028 |
| Kenya | AFR | LMIC | Gavi | Yes | Yes | No | 2032 |
| Kyrgyz Republic | EUR | LMIC | Gavi | No | No | Yes | 2044 |
| Cambodia | WPR | LMIC | Gavi | No | No | No | 2036 |
| Lao People’s Democratic Republic | WPR | LMIC | Gavi | No | No | No | 2035 |
| Liberia | AFR | LIC | Gavi | Yes | Yes | No | 2037 |
| Libya | EMR | UMIC | Non-Gavi | No | No | No | 2035 |
| Sri Lanka | SEAR | LMIC | Gavi | No | No | No | 2028 |
| Lesotho | AFR | LMIC | Gavi | Yes | Yes | No | 2039 |
| Morocco | EMR | LMIC | Non-Gavi | No | No | No | 2029 |
| Moldova, Republic of | EUR | LMIC | Gavi | No | No | Yes | 2034 |
| Madagascar | AFR | LIC | Gavi | No | No | No | 2031 |
| Maldives | SEAR | UMIC | Non-Gavi | No | No | No | 2034 |
| Mexico | AMR | UMIC | Non-Gavi | No | No | No | 2029 |
| Mali | AFR | LIC | Gavi | No | No | No | 2037 |
| Myanmar | SEAR | LMIC | Gavi | Yes | Yes | Yes | 2031 |
| Montenegro | EUR | UMIC | Non-Gavi | No | No | No | 2044 |
| Mongolia | WPR | LMIC | Gavi | Yes | No | Yes | 2032 |

| | | | | | | | |
|---|---|---|---|---|---|---|---|
| Mozambique | AFR | LIC | Gavi | Yes | Yes | Yes | 2032 |
| Mauritania | AFR | LMIC | Gavi | No | No | No | 2042 |
| Malawi | AFR | LIC | Gavi | No | Yes | No | 2038 |
| Malaysia | WPR | UMIC | Non-Gavi | No | No | No | 2028 |
| Namibia | AFR | UMIC | Non-Gavi | Yes | Yes | No | 2030 |
| Niger | AFR | LIC | Gavi | No | No | No | 2036 |
| Nigeria | AFR | LMIC | Gavi | Yes | Yes | Yes | 2030 |
| Nicaragua | AMR | LMIC | Gavi | No | No | No | 2047 |
| Nepal | SEAR | LMIC | Gavi | No | No | Yes | 2036 |
| Pakistan | EMR | LMIC | Gavi | Yes | No | Yes | 2031 |
| Peru | AMR | UMIC | Non-Gavi | No | No | Yes | 2029 |
| Philippines | WPR | LMIC | Non-Gavi | Yes | Yes | Yes | 2030 |
| Papua New Guinea | WPR | LMIC | Gavi | Yes | No | Yes | 2032 |
| Paraguay | AMR | UMIC | Non-Gavi | No | No | No | 2035 |
| Russian Federation | EUR | UMIC | Non-Gavi | No | Yes | Yes | 2030 |
| Rwanda | AFR | LIC | Gavi | No | No | No | 2045 |
| Sudan | EMR | LIC | Gavi | No | No | No | 2036 |
| Senegal | AFR | LMIC | Gavi | No | No | No | 2038 |
| Solomon Islands | WPR | LMIC | Gavi | No | No | No | 2047 |
| Sierra Leone | AFR | LIC | Gavi | Yes | No | No | 2037 |
| El Salvador | AMR | LMIC | Non-Gavi | No | No | No | 2039 |
| Serbia | EUR | UMIC | Non-Gavi | No | No | No | 2036 |
| South Sudan | AFR | LIC | Gavi | No | No | No | 2034 |
| São Tomé and Principe | AFR | LMIC | Gavi | No | No | No | 2044 |
| Suriname | AMR | UMIC | Non-Gavi | No | No | No | 2040 |
| Swaziland | AFR | LMIC | Non-Gavi | No | Yes | No | 2036 |
| Syrian Arab Republic | EMR | LIC | Gavi | No | No | No | 2036 |
| Chad | AFR | LIC | Gavi | No | No | No | 2033 |
| Togo | AFR | LIC | Gavi | No | No | No | 2041 |
| Thailand | SEAR | UMIC | Non-Gavi | Yes | Yes | No | 2031 |
| Tajikistan | EUR | LIC | Gavi | No | No | Yes | 2045 |
| Timor-Leste | SEAR | LMIC | Gavi | No | No | No | 2031 |

| | | | | | | | |
|---|---|---|---|---|---|---|---|
| Tunisia | EMR | LMIC | Non-Gavi | No | No | No | 2036 |
| Turkey | EUR | UMIC | Non-Gavi | No | No | No | 2030 |
| Tanzania, United Republic of | AFR | LMIC | Gavi | Yes | Yes | No | 2031 |
| Uganda | AFR | LIC | Gavi | Yes | Yes | No | 2034 |
| Ukraine | EUR | LMIC | Non-Gavi | No | No | Yes | 2033 |
| Uzbekistan | EUR | LMIC | Gavi | No | No | Yes | 2038 |
| Venezuela | AMR | UMIC | Non-Gavi | No | No | No | 2035 |
| Vietnam | WPR | LMIC | Gavi | Yes | No | Yes | 2038 |
| Vanuatu | WPR | LMIC | Non-Gavi | No | No | No | 2042 |
| Yemen | EMR | LIC | Gavi | No | No | No | 2036 |
| South Africa | AFR | UMIC | Non-Gavi | Yes | Yes | Yes | 2029 |
| Zambia | AFR | LIC | Gavi | Yes | Yes | Yes | 2034 |
| Zimbabwe | AFR | LMIC | Gavi | No | Yes | Yes | 2032 |

[a] LIC: Gross national income (GNI) per capita of $1,085 or less; LMIC: GNI per capita of $1,086 to $4,225; UMIC: GNI per capita of $4,256 to $13,205 (World Bank 2021).

[b] High-TB, high-TB/HIV (HIV-associated TB), and high-MDR/RR-TB (multidrug/rifampicin-resistant TB) burden countries as defined by the World Health Organization [19].

Note: Vaccine introduction was assumed to commence in 2028 and end in 2047. See Clark et al. (https://doi.org/10.1101/2022.04.16.22273762) for introduction year methodology. AFR = African region; AMR = Region of the Americas; EMR = Eastern Mediterranean region; EUR = European region; LIC = low-income; LMIC = lower middle-income; SEAR = Southeast Asian region; TB = tuberculosis; UMIC = upper middle-income; WPR = Western Pacific region.

### S3. Disability weight assumptions: mean, lower bound, upper bound [20].

| Disability weight | Mean | Lower bound | Upper bound |
|---|---|---|---|
| TB | 0.333 | 0.224 | 0.454 |
| TB/HIV+ | 0.408 | 0.274 | 0.549 |
| HIV+ | 0.2893 | 0.1987 | 0.3837 |
| ART | 0.078 | 0.052 | 0.111 |

Note: ART = antiretroviral therapy; HIV = human immunodeficiency virus; TB = tuberculosis.

### S4. The CHEERS 2022 checklist

**From: Consolidated Health Economic Evaluation Reporting Standards 2022 (CHEERS 2022) statement: updated reporting guidance for health economic evaluations [21].**

| Section/topic | Item No | Guidance for reporting | Reported in section |
|---|---|---|---|
| **Title** | | | |
| Title | 1 | Identify the study as an economic evaluation and specify the interventions being compared. | Title Page, Paragraph 1 |
| **Abstract** | | | |
| Abstract | 2 | Provide a structured summary that highlights context, key methods, results, and alternative analyses. | Abstract, Paragraphs 1–4 |
| **Introduction** | | | |
| Background and objectives | 3 | Give the context for the study, the study question, and its practical relevance for decision making in policy or practice. | Background, Paragraphs 1–3 |
| **Methods** | | | |
| Health economic analysis plan | 4 | Indicate whether a health economic analysis plan was developed and where available. | Methods, Analytic overview, Paragraph 1 |
| Study population | 5 | Describe characteristics of the study population (such as age range, demographics, socioeconomic, or clinical characteristics). | Methods, Vaccination scenarios, Paragraph 1<br>Appendix S1, section S1.5, Paragraph 1 |
| Setting and location | 6 | Provide relevant contextual information that may influence findings. | Methods, Analytic overview, Paragraph 1<br>Appendix S2 |
| Comparators | 7 | Describe the interventions or strategies being compared and why chosen. | Methods, Analytic overview, Paragraph 1<br>Methods, Vaccination scenarios, Paragraph 1 |
| Perspective | 8 | State the perspective(s) adopted by the study and why chosen. | Methods, Cost-effectiveness analysis, Paragraph 1_______ |

| Section/topic | Item No | Guidance for reporting | Reported in section |
|---|---|---|---|
| Time horizon | 9 | State the time horizon for the study and why appropriate. | Methods, Vaccination scenarios, Paragraph 1<br>Appendix S1, section S1.5, Paragraph 1 |
| Discount rate | 10 | Report the discount rate(s) and reason chosen. | Methods, Cost-effectiveness analysis, Paragraph 1 |
| Selection of outcomes | 11 | Describe what outcomes were used as the measure(s) of benefit(s) and harm(s). | Methods, Summary health outcomes, Paragraph 1 |
| Measurement of outcomes | 12 | Describe how outcomes used to capture benefit(s) and harm(s) were measured. | Methods, Cost outcomes, Paragraphs 1–4 |
| Valuation of outcomes | 13 | Describe the population and methods used to measure and value outcomes. | Methods, Summary health outcomes, Paragraph 1<br>Methods, Cost outcomes, Paragraphs 1–4 |
| Measurement and valuation of resources and costs | 14 | Describe how costs were valued. | Methods, Cost outcomes, Paragraphs 1–4 |
| Currency, price date, and conversion | 15 | Report the dates of the estimated resource quantities and unit costs, plus the currency and year of conversion. | Methods, Cost outcomes, Paragraphs 1–4 |
| Rationale and description of model | 16 | If modelling is used, describe in detail and why used. Report if the model is publicly available and where it can be accessed. | Appendix S1 [1] |
| Analytics and assumptions | 17 | Describe any methods for analysing or statistically transforming data, any extrapolation methods, and approaches for validating any model used. | Methods, Cost outcomes, Paragraphs 1–4 |
| Characterising heterogeneity | 18 | Describe any methods used for estimating how the results of the study vary for subgroups. | Not applicable |
| Characterising distributional effects | 19 | Describe how impacts are distributed across different individuals or adjustments made to reflect priority populations. | Not applicable |
| Characterising uncertainty | 20 | Describe methods to characterise any sources of uncertainty in the analysis. | Methods, Statistical analysis, Paragraph 1<br>Methods, Sensitivity analysis, Paragraphs 1–7 |

| Section/topic | Item No | Guidance for reporting | Reported in section |
|---|---|---|---|
| Approach to engagement with patients and others affected by the study | 21 | Describe any approaches to engage patients or service recipients, the general public, communities, or stakeholders (such as clinicians or payers) in the design of the study. | Not applicable |
| **Results** | | | |
| Study parameters | 22 | Report all analytic inputs (such as values, ranges, references) including uncertainty or distributional assumptions. | Results, Table 1<br>Appendix S3 |
| Summary of main results | 23 | Report the mean values for the main categories of costs and outcomes of interest and summarise them in the most appropriate overall measure. | Results, Costs and cost-effectiveness, Paragraphs 1–5 |
| Effect of uncertainty | 24 | Describe how uncertainty about analytic judgments, inputs, or projections affect findings. Report the effect of choice of discount rate and time horizon, if applicable. | Results, Sensitivity analysis, Paragraphs 1–7 |
| Effect of engagement with patients and others affected by the study | 25 | Report on any difference patient/service recipient, general public, community, or stakeholder involvement made to the approach or findings of the study | Not applicable |
| **Discussion** | | | |
| Study findings, limitations, generalisability, and current knowledge | 26 | Report key findings, limitations, ethical or equity considerations not captured, and how these could affect patients, policy, or practice. | Discussion, Paragraphs 1–6 |
| **Other relevant information** | | | |
| Source of funding | 27 | Describe how the study was funded and any role of the funder in the identification, design, conduct, and reporting of the analysis | Acknowledgments, Paragraph 1 |
| Conflicts of interest | 28 | Report authors conflicts of interest according to journal or International Committee of Medical Journal Editors requirements. | Supplementary material |

### S5. Undiscounted total health system costs (billions) of no-vaccine scenario by programmatic area across 2028–2050.

| Country grouping | Drug-susceptible TB direct medical costs | Rifampicin-resistant TB direct medical costs | ART costs | Vaccination costs |
|---|---|---|---|---|
| All countries | 20.7 (12.8–31.2) | 19.2 (15.6–23.1) | 397 (340–445) | 0 |
| High-TB burden[a] | 18.7 (11.5328.2) | 15.8 (12.9–19.0) | 161 (140–179) | 0 |
| High-TB/HIV burden[a] | 14.6 (8.96–22.1) | 12.5 (10.2–15.1) | 172 (149–193) | 0 |
| High-MDR/RR-TB burden[a] | 17.4 (10.7–26.2) | 17.7 (14.4–21.3) | 158 (138–176) | 0 |
| **Income level[b]** | | | | |
| LIC | 1.30 (0.83–1.90) | 0.67 (0.52–0.84) | 83.2 (70.6–93.8) | 0 |
| LMIC | 12.5 (7.73–18.9) | 11.3 (9.07–13.8) | 177 (149–199) | 0 |
| UMIC | 6.89 (4.2–10.4) | 7.18 (6.03–8.41) | 137 (120–152) | 0 |
| **World region** | | | | |
| AFR | 4.23 (2.59–6.34) | 2.96 (2.38–3.60) | 198 (170–223) | 0 |
| AMR | 0.61 (0.38–0.92) | 0.35 (0.29–0.42) | 38.0 (33.2–42.2) | 0 |
| EMR | 1.28 (0.80–1.90) | 1.49 (1.18–1.82) | 35.5 (30.1–39.7) | 0 |
| EUR | 0.29 (0.18–0.43) | 2.28 (1.87–2.74) | 42.9 (36.9–47.5) | 0 |
| SEAR | 10.2 (6.25–15.5) | 8.17 (6.61–9.84) | 19.4 (15.8–22.2) | 0 |
| WPR | 4.09 (2.58–6.04) | 3.94 (3.30–4.64) | 63.3 (54.0–70.2) | 0 |

[a] High-TB, high-TB/HIV (HIV-associated TB), and high-MDR/RR-TB (multidrug/rifampicin-resistant TB) burden countries as defined by the World Health Organization [19].

[b] LIC: Gross national income (GNI) per capita of $1,085 or less; LMIC: GNI per capita of $1,086 to $4,225; UMIC: GNI per capita of $4,256 to $13,205 (World Bank 2021).

Note: All countries include 105 low- and middle-income countries analyzed. Values in parentheses represent equal-tailed 95% credible intervals. AFR = African region; AMR = Region of the Americas; EMR = Eastern Mediterranean region; EUR = European region; LIC = low-income; LMIC = lower middle-income; SEAR = Southeast Asian region; TB = tuberculosis; UMIC = upper middle-income; WPR = Western Pacific region.

### S6. Undiscounted total health system costs (billions) of infant tuberculosis vaccines by programmatic area across 2028–2050 and percent change compared to no vaccination.

| Country grouping | Drug-susceptible TB direct medical costs | Rifampicin-resistant TB direct medical costs | ART costs | Vaccination costs |
|---|---|---|---|---|
| All countries | 20.3 (12.5–30.7)<br>*1.7% reduction* | 18.9 (15.4–22.7)<br>*1.6% reduction* | 397 (340–445)<br>*0.01% increase* | 11.8 (9.59–16.9) |
| High-TB burden[a] | 18.4 (11.3–27.7)<br>*1.7% reduction* | 15.6 (12.7–18.7)<br>*1.8% reduction* | 161 (140–179)<br>*0.01% increase* | 7.99 (6.65–11.2) |
| High-TB/HIV burden[a] | 14.3 (8.78–21.7)<br>*1.8% reduction* | 12.3 (9.97–14.8)<br>*1.7% reduction* | 172 (149–193)<br>*0.01% increase* | 5.71 (4.8–7.81) |
| High-MDR/RR-TB burden[a] | 17.1 (10.5–25.8)<br>*1.7% reduction* | 17.4 (14.2–20.9)<br>*1.6% reduction* | 158 (138–176)<br>*0.01% increase* | 7.06 (5.85–10.0) |
| **Income level[b]** | | | | |
| LIC | 1.27 (0.81–1.85)<br>*2.4% reduction* | 0.65 (0.51–0.82)<br>*2.3% reduction* | 83.2 (70.6–93.8)<br>*0.005% increase* | 1.84 (1.57–2.41) |
| LMIC | 12.3 (7.58–18.5)<br>*1.8% reduction* | 11.1 (8.89–13.6)<br>*1.9% reduction* | 177 (149–199)<br>*0.01% increase* | 5.68 (4.80–7.65) |
| UMIC | 6.81 (4.15–10.27)<br>*1.2% reduction* | 7.12 (5.98–8.33)<br>*0.9% reduction* | 137 (120–152)<br>*0.01% increase* | 4.26 (3.22–6.80) |
| **World region** | | | | |
| AFR | 4.12 (2.52–6.20)<br>*2.5% reduction* | 2.88 (2.31–3.51)<br>*2.6% reduction* | 198 (170–223)<br>*0.01% increase* | 3.73 (3.16–4.99) |
| AMR | 0.61 (0.37–0.91)<br>*0.5% reduction* | 0.35 (0.28–0.42)<br>*0.7% reduction* | 38.0 (33.2–42.2)<br>*0.01% increase* | 0.99 (0.75–1.56) |
| EMR | 1.24 (0.77–1.85)<br>*3.1% reduction* | 1.44 (1.14–1.77)<br>*3.3% reduction* | 35.5 (30.1–39.7)<br>*0.01% increase* | 1.64 (1.35–2.25) |
| EUR | 0.29 (0.18–0.43)<br>*0.4% reduction* | 2.27 (1.86–2.74)<br>*0.4% reduction* | 42.9 (36.9–47.5)<br>*0.005% increase* | 0.63 (0.44–1.07) |
| SEAR | 10.1 (6.16–15.31)<br>*1.3% reduction* | 8.04 (6.51–9.70)<br>*1.5% reduction* | 19.4 (15.8–22.2)<br>*0.01% increase* | 2.6 (2.18–3.54) |
| WPR | 4.03 (2.54–5.96)<br>*1.4% reduction* | 3.90 (3.27–4.60)<br>*1.0% reduction* | 63.3 (54.0–70.3)<br>*0.01% increase* | 2.19 (1.71–3.44) |

[a] High-TB, high-TB/HIV (HIV-associated TB), and high-MDR/RR-TB (multidrug/rifampicin-resistant TB) burden countries as defined by the World Health Organization [19].

[b] LIC: Gross national income (GNI) per capita of $1,085 or less; LMIC: GNI per capita of $1,086 to $4,225; UMIC: GNI per capita of $4,256 to $13,205 (World Bank 2021).

Note: All countries include 105 low- and middle-income countries analyzed. Values in parentheses represent equal-tailed 95% credible intervals. AFR = African region; AMR = Region of the Americas; EMR = Eastern Mediterranean region; EUR = European region; LIC = low-income; LMIC = lower middle-income; SEAR = Southeast Asian region; TB = tuberculosis; UMIC = upper middle-income; WPR = Western Pacific region.

**S7. Undiscounted total health system costs (billions) of adolescent/adult tuberculosis vaccines by programmatic area across 2028–2050 and percent change compared to no vaccination.**

| Country grouping | Drug-susceptible TB direct medical costs | Rifampicin-resistant TB direct medical costs | ART costs | Vaccination costs |
|---|---|---|---|---|
| All countries | 17.2 (10.6–25.9)<br>*16.8% reduction* | 16.0 (13.0–19.3)<br>*16.4% reduction* | 398 (340–446)<br>*0.21% increase* | 50.5 (38.1–75.9) |
| High-TB burden[a] | 15.5 (9.54–23.4)<br>*17.1% reduction* | 13.1 (10.6–15.8)<br>*17.2% reduction* | 161 (140–180)<br>*0.27% increase* | 36.6 (27.7–54.7) |
| High-TB/HIV burden[a] | 12.0 (7.37–18.2)<br>*17.5% reduction* | 10.4 (8.38–12.5)<br>*17.1% reduction* | 173 (149–193)<br>*0.25% increase* | 23.5 (18.4–33.3) |
| High-MDR/RR-TB burden[a] | 14.4 (8.89–21.8)<br>*16.9% reduction* | 14.8 (12.0–17.8)<br>*16.5% reduction* | 158 (139–176)<br>*0.26% increase* | 34.1 (25.8–51.4) |
| **Income level[b]** | | | | |
| LIC | 1.06 (0.68–1.55)<br>*17.9% reduction* | 0.56 (0.44–0.70)<br>*16.4% reduction* | 83.3 (70.7–93.9)<br>*0.15% increase* | 5.60 (4.50–7.46) |
| LMIC | 10.4 (6.41–15.7)<br>*17.0% reduction* | 9.41 (7.49–11.5)<br>*17.0% reduction* | 178 (150–200)<br>*0.19% increase* | 23.1 (18.6–31.1) |
| UMIC | 5.77 (3.51–8.72)<br>*16.3% reduction* | 6.07 (5.07–7.12)<br>*15.5% reduction* | 137 (120–152)<br>*0.26% increase* | 21.9 (15.0–37.4) |
| **World region** | | | | |
| AFR | 3.44 (2.10–5.20)<br>*18.5% reduction* | 2.41 (1.91–2.95)<br>*18.6% reduction* | 199 (170–224)<br>*0.23% increase* | 9.96 (7.98–13.5) |
| AMR | 0.53 (0.32–0.79)<br>*13.7% reduction* | 0.30 (0.24–0.36)<br>*14.8% reduction* | 38.0 (33.2–42.3)<br>*0.18% increase* | 4.78 (3.29–8.17) |
| EMR | 1.04 (0.64–1.55)<br>*18.9% reduction* | 1.20 (0.94–1.47)<br>*19.5% reduction* | 35.6 (30.2–39.8)<br>*0.17% increase* | 5.66 (4.49–7.86) |
| EUR | 0.25 (0.16–0.38)<br>*12.7% reduction* | 2.00 (1.64–2.42)<br>*12.2% reduction* | 43.0 (37.0–47.5)<br>*0.16% increase* | 2.82 (1.98–4.72) |
| SEAR | 8.52 (5.22–13.0)<br>*16.5% reduction* | 6.79 (5.50–8.16)<br>*16.8% reduction* | 19.5 (15.9–22.2)<br>*0.22% increase* | 14.6 (11.7–20) |
| WPR | 3.43 (2.16–5.08)<br>*16.1% reduction* | 3.33 (2.77–3.95)<br>*15.3% reduction* | 63.4 (54.1–70.4)<br>*0.19% increase* | 12.7 (8.73–21.7) |

[a] High-TB, high-TB/HIV (HIV-associated TB), and high-MDR/RR-TB (multidrug/rifampicin-resistant TB) burden countries as defined by the World Health Organization [19].

[b] LIC: Gross national income (GNI) per capita of $1,085 or less; LMIC: GNI per capita of $1,086 to $4,225; UMIC: GNI per capita of $4,256 to $13,205 (World Bank 2021).

Note: All countries include 105 low- and middle-income countries analyzed. Values in parentheses represent equal-tailed 95% credible intervals. AFR = African region; AMR = Region of the Americas; EMR = Eastern Mediterranean region; EUR = European region; LIC = low-income; LMIC = lower middle-income; SEAR = Southeast Asian region; TB = tuberculosis; UMIC = upper middle-income; WPR = Western Pacific region.

### S8. Undiscounted incremental societal costs (millions) of infant tuberculosis vaccines across 2028–2050 and percent change compared to no vaccination.

| Country grouping | Drug-susceptible TB non-medical costs | Rifampicin-resistant TB non-medical costs | Drug-susceptible TB productivity loss due to testing and treatment[a] | Rifampicin-resistant TB productivity loss due to testing and treatment[a] | Productivity loss due to premature death |
|---|---|---|---|---|---|
| All countries | 772 (673–897) | 100 (91.8–114) | 855 (740–986) | 93.4 (80.5–109) | 57200 (49700–65300) |
| | *1.7% reduction* | *1.3% reduction* | *1.8% reduction* | *1.5% reduction* | *0.1% reduction* |
| High-TB burden[b] | 667 (581–776) | 82.7 (76.0–94.0) | 754 (651–861) | 80.8 (69.1–93.8) | 53800 (46300–61900) |
| | *1.7% reduction* | *1.6% reduction* | *1.8% reduction* | *1.7% reduction* | *0.1% reduction* |
| High-TB/HIV burden[b] | 546 (477–625) | 64.6 (58.7–72.4) | 653 (565–739) | 68.6 (58.3–80.8) | 46400 (39300–54600) |
| | *1.9% reduction* | *1.6% reduction* | *1.9% reduction* | *1.8% reduction* | *0.2% reduction* |
| High-MDR/RR-TB burden[b] | 597 (518–695) | 85.3 (78.5–96.6) | 678 (588–774) | 80.6 (69.2–93.6) | 50300 (42900–58400) |
| | *1.7% reduction* | *1.2% reduction* | *1.8% reduction* | *1.5% reduction* | *0.1% reduction* |
| **Income level[c]** | | | | | |
| LIC | 144 (122–170) | 15.0 (12.9–18.0) | 127 (106–159) | 10.8 (8.96–13.8) | 116100 (115800–116400) |
| | *2.9% reduction* | *2.6% reduction* | *3% reduction* | *3% reduction* | *7.7% reduction* |
| LMIC | 482 (421–560) | 58.0 (53.9–64.8) | 469 (417–535) | 44.9 (39.8–51.7) | 44300 (37400–52100) |
| | *1.9% reduction* | *1.8% reduction* | *2% reduction* | *1.9% reduction* | *0.2% reduction* |
| UMIC | 146 (131–167) | 27.3 (25.0–31.7) | 260 (217–292) | 37.7 (31.7–43.5) | 10600 (8450–13600) |
| | *1.1% reduction* | *0.7% reduction* | *1.3% reduction* | *1.1% reduction* | *0.02% reduction* |
| **World region** | | | | | |
| AFR | 259 (223–297) | 30.0 (26.7–34.0) | 337 (282–373) | 39.6 (33.9–46.3) | 25600 (20700–31600) |
| | *2.4% reduction* | *2.4% reduction* | *2.3% reduction* | *2.3% reduction* | *0.3% reduction* |
| AMR | 8.19 (6.93–9.39) | 1.27 (1.08–1.36) | 14.0 (12.7–15.0) | 1.53 (1.40–1.75) | 617 (524–732) |
| | *0.5% reduction* | *0.7% reduction* | *0.5% reduction* | *0.6% reduction* | *0.01% reduction* |
| EMR | 139 (119–170) | 19.8 (17.7–23.6) | 122 (110–152) | 14.0 (12.7–16.4) | 117000 (116000–118000) |
| | *3.7% reduction* | *3.7% reduction* | *3.7% reduction* | *3.7% reduction* | *3.6% reduction* |
| EUR | 4.20 (3.60–4.64) | 6.00 (5.42–6.74) | 3.22 (2.86–3.78) | 3.52 (3.34–3.87) | 349 (297–406) |
| | *0.4% reduction* | *0.4% reduction* | *0.4% reduction* | *0.4% reduction* | *0% reduction* |
| SEAR | 244 (207–296) | 28.9 (26.3–33.2) | 236 (198–286) | 22.2 (18.0–27.2) | 19000 (15000–24100) |
| | *1.4% reduction* | *1.6% reduction* | *1.4% reduction* | *1.6% reduction* | *0.1% reduction* |
| WPR | 117 (113–120) | 14.3 (14.6–15.5) | 142 (134–155) | 12.6 (11.2–13.7) | 8690 (6720–11400) |
| | *1.2% reduction* | *0.7% reduction* | *1.5% reduction* | *0.9% reduction* | *0.02% reduction* |

[a] Costs include patient costs due to time lost for tuberculosis testing and treatment.
[b] High-TB, high-TB/HIV (HIV-associated TB), and high-MDR/RR-TB (multidrug/rifampicin-resistant TB) burden countries as defined by the World Health Organization [19].

[c] LIC: Gross national income (GNI) per capita of $1,085 or less; LMIC: GNI per capita of $1,086 to $4,225; UMIC: GNI per capita of $4,256 to $13,205 (World Bank 2021).

Note: All countries include 105 low- and middle-income countries analyzed. Values in parentheses represent equal-tailed 95% credible intervals. AFR = African region; AMR = Region of the Americas; EMR = Eastern Mediterranean region; EUR = European region; LIC = low-income; LMIC = lower middle-income; SEAR = Southeast Asian region; TB = tuberculosis; UMIC = upper middle-income; WPR = Western Pacific region.

**S9. Undiscounted incremental societal costs (billions) of adolescent/adult tuberculosis vaccines across 2028–2050 and percent change compared to no vaccination.**

| Country grouping | Drug-susceptible TB non-medical costs | Rifampicin-resistant TB non-medical costs | Drug-susceptible TB productivity loss due to testing and treatment[a] | Rifampicin-resistant TB productivity loss due to testing and treatment[a] | Productivity loss due to premature death |
|---|---|---|---|---|---|
| All countries | 7.45 (6.39–8.62)<br>*16.8% reduction* | 1.18 (1.02–1.35)<br>*15.6% reduction* | 8.09 (6.91–9.29)<br>*17% reduction* | 0.99 (0.85–1.15)<br>*16.2% reduction* | 376 (354–399)<br>*0.5% reduction* |
| High-TB burden[b] | 6.52 (5.6–7.53)<br>*17.0% reduction* | 0.89 (0.77–1.01)<br>*16.6% reduction* | 7.13 (6.1–8.15)<br>*17.2% reduction* | 0.80 (0.68–0.91)<br>*17.1% reduction* | 243 (221–266)<br>*0.4% reduction* |
| High-TB/HIV burden[b] | 5.10 (4.36–5.90)<br>*17.5% reduction* | 0.66 (0.56–0.76)<br>*16.6% reduction* | 6.01 (5.12–6.87)<br>*17.6% reduction* | 0.66 (0.56–0.76)<br>*17.1% reduction* | 215 (194–239)<br>*0.8% reduction* |
| High-MDR/RR-TB burden[b] | 6.00 (5.15–6.94)<br>*16.8% reduction* | 1.06 (0.92–1.21)<br>*15.5% reduction* | 6.44 (5.51–7.37)<br>*17.2% reduction* | 0.88 (0.75–1.01)<br>*16.3% reduction* | 228 (206–252)<br>*0.4% reduction* |
| **Income level[c]** | | | | | |
| LIC | 0.91 (0.76–1.08)<br>*18.3% reduction* | 0.09 (0.07–0.11)<br>*15.6% reduction* | 0.78 (0.65–0.93)<br>*18.5% reduction* | 0.06 (0.05–0.08)<br>*17.0% reduction* | 122 (121–123)<br>*8.1% reduction* |
| LMIC | 4.40 (3.75–5.16)<br>*16.9% reduction* | 0.53 (0.45–0.62)<br>*16.1% reduction* | 4.11 (3.47–4.74)<br>*17.1% reduction* | 0.39 (0.33–0.46)<br>*16.5% reduction* | 202 (183–223)<br>*1% reduction* |
| UMIC | 2.14 (1.89–2.39)<br>*16% reduction* | 0.56 (0.49–0.62)<br>*15.0% reduction* | 3.20 (2.79–3.62)<br>*16.5% reduction* | 0.54 (0.46–0.61)<br>*15.9% reduction* | 51.9 (43.7–63.2)<br>*0.1% reduction* |
| **World region** | | | | | |
| AFR | 1.97 (1.68–2.26)<br>*18% reduction* | 0.22 (0.19–0.26)<br>*17.9% reduction* | 2.65 (2.27–3.05)<br>*17.9% reduction* | 0.31 (0.27–0.36)<br>*17.8% reduction* | 92.3 (80.0–108)<br>*1.25% reduction* |
| AMR | 0.21 (0.18–0.24)<br>*13.8% reduction* | 0.03 (0.02–0.03)<br>*15.0% reduction* | 0.40 (0.34–0.46)<br>*13.5% reduction* | 0.04 (0.03–0.05)<br>*14.1% reduction* | 5.13 (4.67–5.68)<br>*0.05% reduction* |
| EMR | 0.76 (0.65–0.90)<br>*20.2% reduction* | 0.11 (0.09–0.13)<br>*20.2% reduction* | 0.67 (0.57–0.79)<br>*20.1% reduction* | 0.08 (0.07–0.09)<br>*20.3% reduction* | 123 (121–125)<br>*3.8% reduction* |
| EUR | 0.12 (0.10–0.14)<br>*11.9% reduction* | 0.20 (0.17–0.23)<br>*11.8% reduction* | 0.09 (0.08–0.11)<br>*12.1% reduction* | 0.12 (0.10–0.14)<br>*12.1% reduction* | 3.35 (2.96–3.73)<br>*0.04% reduction* |
| SEAR | 2.88 (2.42–3.38)<br>*16.6% reduction* | 0.31 (0.26–0.35)<br>*17.0% reduction* | 2.74 (2.29–3.16)<br>*16.7% reduction* | 0.23 (0.20–0.27)<br>*17.2% reduction* | 121 (104–138)<br>*0.9% reduction* |
| WPR | 1.51 (1.36–1.70)<br>*15.5% reduction* | 0.32 (0.29–0.35)<br>*14.8% reduction* | 1.53 (1.35–1.72)<br>*16.3% reduction* | 0.215 (0.19–0.241)<br>*15.3% reduction* | 31.4 (28.1–35.0)<br>*0.1% reduction* |

[a] Costs include patient costs due to time lost for tuberculosis testing and treatment.

[b] High-TB, high-TB/HIV (HIV-associated TB), and high-MDR/RR-TB (multidrug/rifampicin-resistant TB) burden countries as defined by the World Health Organization [19].

[c] LIC: Gross national income (GNI) per capita of $1,085 or less; LMIC: GNI per capita of $1,086 to $4,225; UMIC: GNI per capita of $4,256 to $13,205 (World Bank 2021).

Note: All countries include 105 low- and middle-income countries analyzed. Values in parentheses represent equal-tailed 95% credible intervals. AFR = African region; AMR = Region of the Americas; EMR = Eastern Mediterranean region; EUR = European region; LIC = low-income; LMIC = lower middle-income; SEAR = Southeast Asian region; TB = tuberculosis; UMIC = upper middle-income; WPR = Western Pacific region.

**S10. Time trend of undiscounted costs (health system perspective) by programmatic area for infant tuberculosis vaccines: (A) including productivity costs; and (B) excluding productivity costs.**

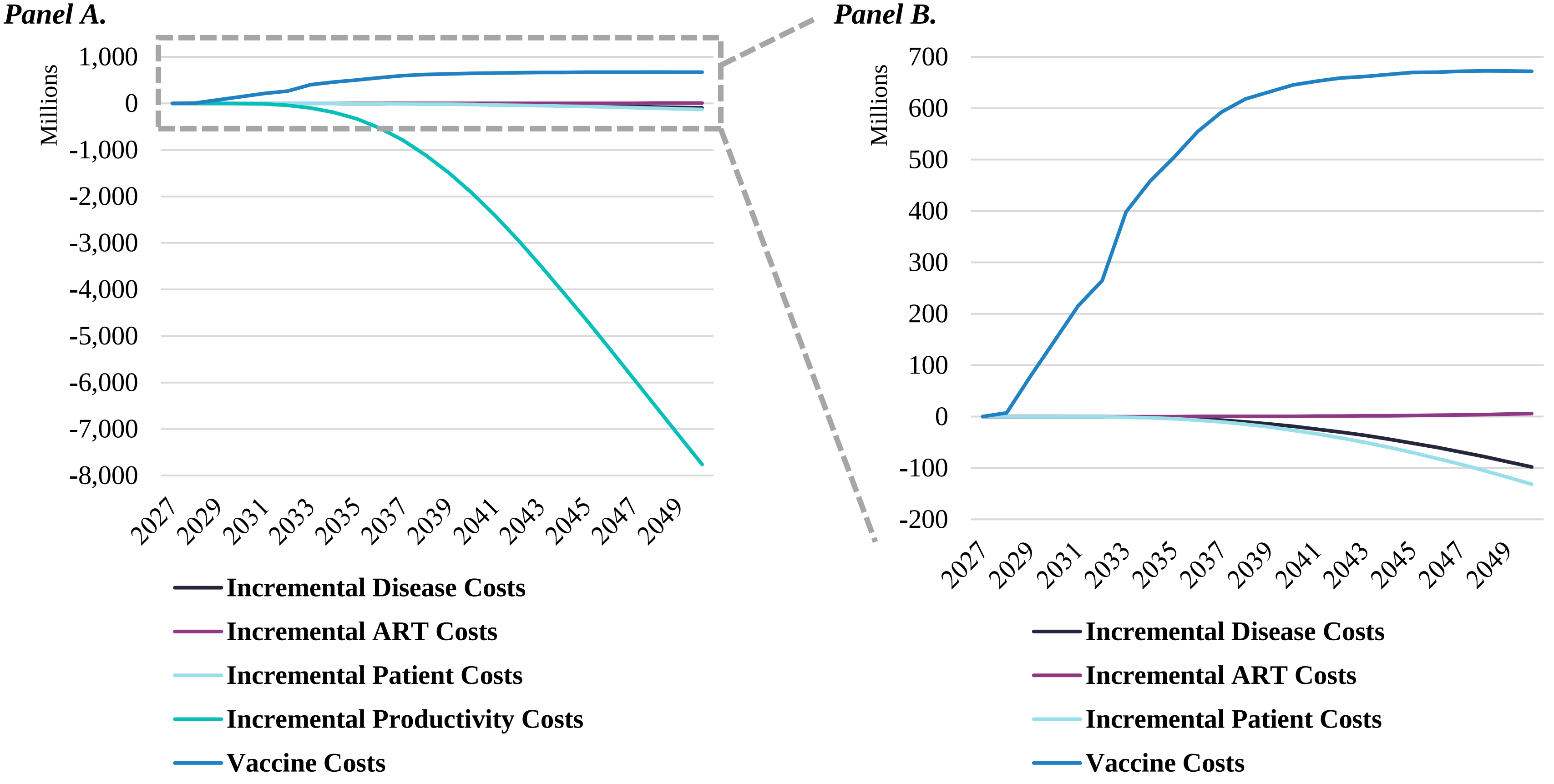


Note: ART = antiretroviral therapy; disease costs = tuberculosis testing and treatment costs from the health system perspective; patient costs = non-medical costs and productivity costs due to time lost for tuberculosis testing and treatment; productivity costs = productivity loss due to premature death.

**S11. Time trend of undiscounted costs (health system perspective) by programmatic area for adolescent/adult tuberculosis vaccines: (A) including productivity costs; and (B) excluding productivity costs.**

***Panel A.*** ***Panel B.***

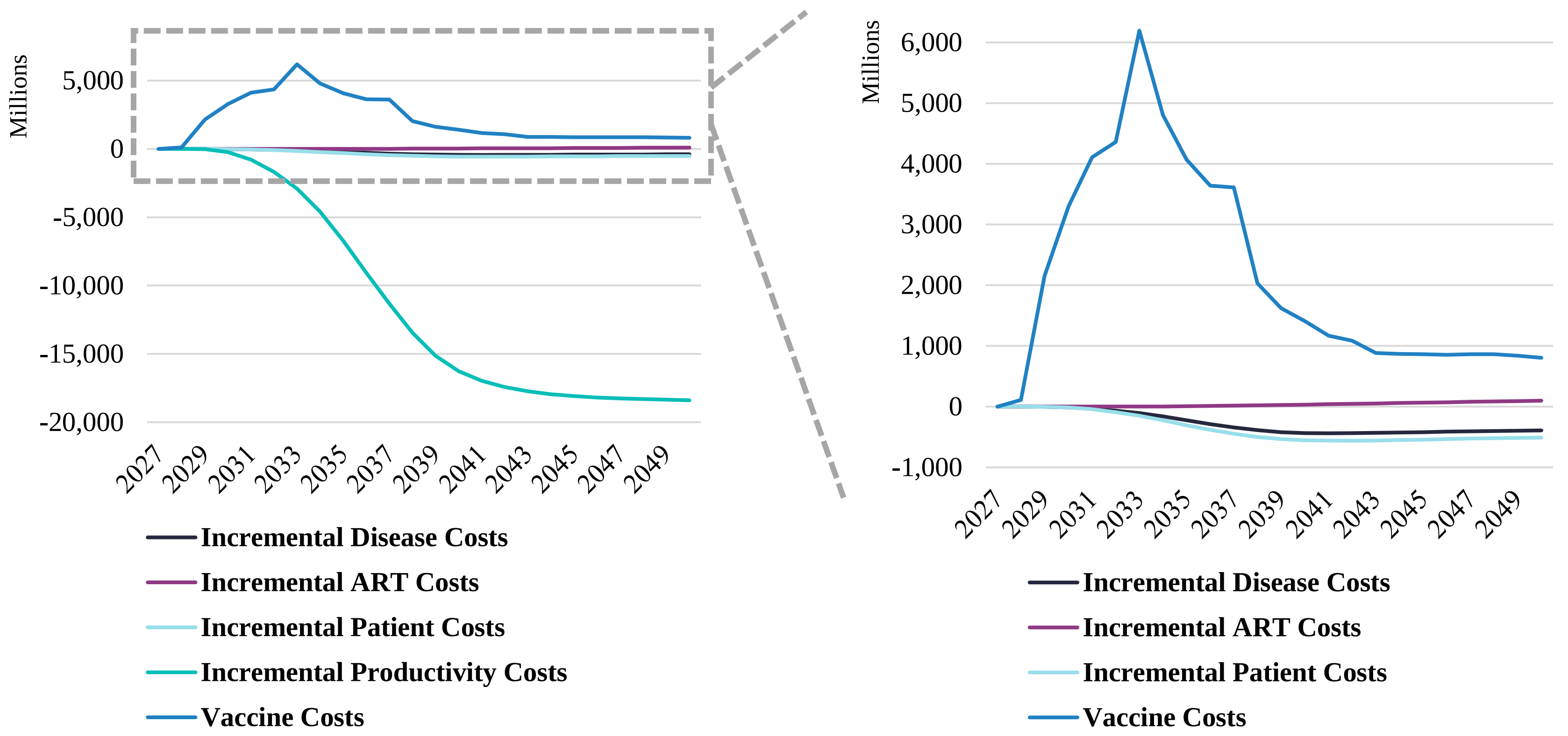


Note: ART = antiretroviral therapy; disease costs = tuberculosis testing and treatment costs from the health system perspective; patient costs = non-medical costs and productivity costs due to time lost for tuberculosis testing and treatment; productivity costs = productivity loss due to premature death.

**S12. Percentage of countries where vaccination was cost-effective compared to percentage of gross domestic product per capita thresholds, comparing health system and societal perspectives.**

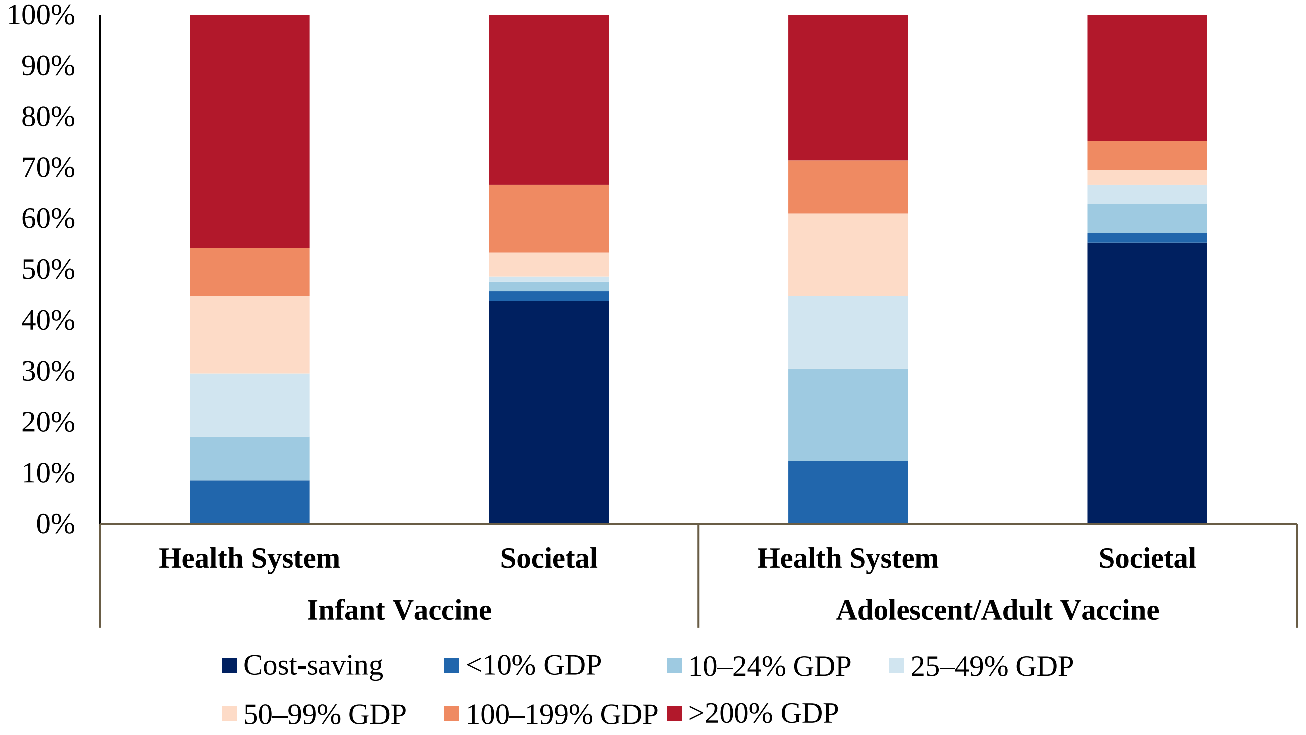


Note: Countries include 105 low- and middle-income countries analyzed. Gross domestic product per capita estimates from 2020. GDP = gross domestic product per capita.

**S13. Percentage of population in countries where vaccination was cost-effective compared to percentage of gross domestic product per capita thresholds, comparing health system and societal perspectives.**

| | Perspective | Cost-saving | <10% GDP | 10–24% GDP | 25–49% GDP | 50–99% GDP | 100–199% GDP | >200% GDP |
|---|---|---|---|---|---|---|---|---|
| Infant Vaccine | Health System | 0% | 11% | 28% | 29% | 9% | 7% | 16% |
| | Societal | 75% | 3% | 2% | 0% | 4% | 6% | 9% |
| Adolescent/ Adult Vaccine | Health System | 0% | 13% | 33% | 32% | 9% | 5% | 9% |
| | Societal | 81% | 4% | 2% | 3% | 1% | 1% | 8% |

Note: Countries include 105 low- and middle-income countries analyzed. Population includes vaccinated individuals 2028–2050. Gross domestic product per capita estimates from 2020. GDP = gross domestic product per capita.

**S14. Percentage of countries that were cost-effective compared to percentage of gross domestic product per capita thresholds, comparing health system and societal perspectives, all countries weighted equally.**

| | Perspective | Cost-saving | <10% GDP | 10–24% GDP | 25–49% GDP | 50–99% GDP | 100–199% GDP | >200% GDP |
|---|---|---|---|---|---|---|---|---|
| Infant Vaccine | Health System | 0% | 9% | 9% | 12% | 15% | 10% | 46% |
| | Societal | 44% | 2% | 2% | 1% | 5% | 13% | 33% |
| Adolescent/ Adult Vaccine | Health System | 0% | 12% | 18% | 14% | 16% | 10% | 29% |
| | Societal | 55% | 2% | 6% | 4% | 3% | 6% | 25% |

Note: Countries include 105 low- and middle-income countries analyzed. Gross domestic product per capita estimates from 2020. GDP = gross domestic product per capita.

### S15. Discounted costs, undiscounted disability-adjusted life-years (DALYs) averted, and cost-effectiveness of infant tuberculosis vaccines.

| Country grouping | Health system perspective[a] incremental cost (USD billions) | Societal perspective[b] incremental cost (USD billions) | DALYs averted (millions) | Health system cost (USD) per DALY averted | Societal cost (USD) per DALY averted |
|---|---|---|---|---|---|
| All countries | 7.67 (6.25, 10.6) | *-27.4* (*-32.6*, *-22.3*) | 74.4 (64.5, 85.9) | 104 (80.4, 146) | cost-saving[c] |
| High-TB burden[d] | 5.15 (4.26, 7.14) | *-28.1* (*-33.1*, *-23.3*) | 68.7 (58.8, 80.3) | 75.4 (58.5, 107.3) | cost-saving[c] |
| High-TB/HIV burden[d] | 3.62 (3.03, 4.88) | *-24.9* (*-29.9*, *-20.5*) | 60.1 (50.6, 71.2) | 60.6 (46.9, 84.1) | cost-saving[c] |
| High-MDR/RR-TB burden[d] | 4.53 (3.72, 6.33) | *-26.5* (*-31.4*, *-21.7*) | 60.6 (50.9, 71.7) | 75.4 (57.4, 109.3) | cost-saving[c] |
| **Income level[e]** | | | | | |
| LIC | 1.12 (0.96, 1.44) | *-0.19* (*-0.48*, 0.15) | 8.41 (7.11, 10.2) | 134 (103, 178) | cost-saving (cost-saving, 13.3) |
| LMIC | 3.95 (3.31, 5.31) | *-23.1* (*-27.8*, *-18.8*) | 61.5 (52.0, 72.9) | 64.7 (50.1, 91.1) | cost-saving[c] |
| UMIC | 2.60 (1.96, 4.12) | *-4.15* (*-6.24*, *-2.25*) | 4.44 (3.36, 5.98) | 599 (373, 974) | cost-saving[c] |
| **World region** | | | | | |
| AFR | 2.40 (2.05, 3.13) | *-13.4* (*-17.1*, *-10.4*) | 38.7 (32.1, 46.6) | 62.8 (47.6, 84.9) | cost-saving[c] |
| AMR | 0.69 (0.52, 1.07) | 0.29 (0.10, 0.67) | 0.25 (0.21, 0.30) | 2730 (1920, 4260) | 1160 (355, 2720) |
| EMR | 1.05 (0.86, 1.42) | *-0.62* (*-1.32*, *-0.01*) | 6.63 (4.58, 9.12) | 164 (105, 250) | cost-saving[c] |
| EUR | 0.43 (0.30, 0.72) | 0.20 (0.07, 0.49) | 0.16 (0.14, 0.18) | 2750 (1860, 4590) | 1300 (419, 3170) |
| SEAR | 1.59 (1.31, 2.18) | *-9.90* (*-12.9*, *-7.37*) | 23.0 (17.2, 30.6) | 70.6 (48.1, 107.9) | cost-saving[c] |
| WPR | 1.50 (1.16, 2.33) | *-4.03* (*-5.74*, *-2.62*) | 5.66 (4.10, 7.91) | 272 (171, 445) | cost-saving[c] |

[a] Costs from the health system perspective include vaccination costs, tuberculosis testing and treatment costs, and antiretroviral treatment costs.
[b] Costs from the societal perspective include health system perspective costs, as well as patient non-medical costs and productivity losses.
[c] Both the point estimate and the interval estimates were cost-saving.
[d] High-TB, high-TB/HIV (HIV-associated TB), and high-MDR/RR-TB (multidrug/rifampicin-resistant TB) burden countries as defined by the World Health Organization [19].
[e] LIC: Gross national income (GNI) per capita of $1,085 or less; LMIC: GNI per capita of $1,086 to $4,225; UMIC: GNI per capita of $4,256 to $13,205 (World Bank 2021).
Note: All countries include 105 low- and middle-income countries analyzed. Values in parentheses represent equal-tailed 95% credible intervals. AFR = African region; AMR = Region of the Americas; EMR = Eastern Mediterranean region; EUR = European region; LIC = low-income; LMIC = lower middle-income; SEAR = Southeast Asian region; UMIC = upper middle-income; USD = United States dollar; WPR = Western Pacific region.

**S16. Discounted costs, undiscounted disability-adjusted life-years (DALYs) averted, and cost-effectiveness of adolescent/adult tuberculosis vaccines.**

| Country grouping | Health system perspective[a] incremental cost (USD billions) | Societal perspective[b] incremental cost (USD billions) | Incremental DALYs averted (millions) | Health system cost (USD) per DALY averted | Societal cost (USD) per DALY averted |
|---|---|---|---|---|---|
| All countries | 36.3 (26.8, 55.2) | *-145* (*-165*, *-123*) | 302 (275, 331) | 120 (86.4, 181) | cost-saving[c] |
| High-TB burden[d] | 26.0 (19.0, 39.8) | *-142* (*-159*, *-122*) | 276 (249, 305) | 94.5 (66.6, 143) | cost-saving[c] |
| High-TB/HIV burden[d] | 15.8 (11.8, 22.9) | *-131* (*-147*, *-115*) | 247 (222, 275) | 64.0 (45.8, 93.2) | cost-saving[c] |
| High-MDR/RR-TB burden[d] | 24.0 (17.2, 37.5) | *-133* (*-151*, *-114*) | 245 (219, 272) | 98.3 (67.4, 152) | cost-saving[c] |
| **Income level[e]** | | | | | |
| LIC | 3.73 (2.98, 4.98) | *-2.57* (*-3.70*, *-1.20*) | 31.4 (27.7, 35.1) | 120 (91.2, 163) | cost-saving[c] |
| LMIC | 17.1 (13.0, 24.3) | *-119* (*-133*, *-105*) | 251 (226, 278) | 68.1 (49.7, 98) | cost-saving[c] |
| UMIC | 15.5 (10.1, 27.4) | *-24.0* (*-33.8*, *-9.93*) | 19.7 (16.0, 25.0) | 796 (479, 1510) | cost-saving[c] |
| **World region** | | | | | |
| AFR | 6.89 (5.51, 9.26) | *-56.6* (*-66.4*, *-48.3*) | 129 (115, 143) | 53.8 (41.4, 74.7) | cost-saving |
| AMR | 3.84 (2.60, 6.56) | *-0.17* (*-1.54*, 2.67) | 1.86 (1.66, 2.05) | 2080 (1380, 3550) | cost-saving (cost-saving, 1450) |
| EMR | 4.06 (3.19, 5.61) | *-2.65* (*-4.45*, *-0.70*) | 21.0 (16.3, 26.3) | 197 (137, 288) | cost-saving |
| EUR | 2.08 (1.40, 3.56) | *-0.64* (*-1.41*, 0.82) | 1.32 (1.17, 1.46) | 1580 (1040, 2790) | cost-saving (cost-saving, 654) |
| SEAR | 9.38 (7.01, 13.6) | *-71.3* (*-82.6*, *-59.8*) | 132 (111, 154) | 71.9 (49.7, 108) | cost-saving |
| WPR | 10.0 (6.55, 17.6) | *-13.8* (*-18.5*, *-5.79*) | 18.1 (15.7, 20.6) | 555 (353, 1000) | cost-saving |

[a] Costs from the health system perspective include vaccination costs, tuberculosis testing and treatment costs, and antiretroviral treatment costs.
[b] Costs from the societal perspective include health system perspective costs, as well as patient non-medical costs and productivity losses.
[c] Both the point estimate and the interval estimates were cost-saving.
[d] High-TB, high-TB/HIV (HIV-associated TB), and high-MDR/RR-TB (multidrug/rifampicin-resistant TB) burden countries as defined by the World Health Organization [19].
[e] LIC: Gross national income (GNI) per capita of $1,085 or less; LMIC: GNI per capita of $1,086 to $4,225; UMIC: GNI per capita of $4,256 to $13,205 (World Bank 2021).
Note: All countries include 105 low- and middle-income countries analyzed. Values in parentheses represent equal-tailed 95% credible intervals. AFR = African region; AMR = Region of the Americas; EMR = Eastern Mediterranean region; EUR = European region; LIC = low-income; LMIC = lower middle-income; SEAR = Southeast Asian region; UMIC = upper middle-income; USD = United States dollar; WPR = Western Pacific region.

**S17. Incremental net monetary benefit (societal perspective, billions) of novel tuberculosis vaccines in countries where vaccine is cost-saving or cost-effective, comparing alternative willingness-to-pay thresholds.**

| Threshold | Infant Vaccine | | | | Adolescent/Adult Vaccine | | | |
|---|---|---|---|---|---|---|---|---|
| | 105 LMICs | Cost-saving | Cost-effective | No. cost-effective | 105 LMICs | Cost-saving | Cost-effective | No. cost-effective |
| 1x per-capita GDP (base-case) | 67.7 (42.6, 99.8) | 68.3 (44.4, 99.5) | 68.6 (44.5, 100) | 56 | 369 (278, 472) | 369 (283, 469) | 372 (283, 474) | 73 |
| 0.5x per-capita GDP | 47.6 (28.7, 71.1) | 48.6 (30.8, 71.4) | 48.7 (30.8, 71.8) | 51 | 257 (187, 333) | 259 (194, 333) | 260 (194, 335) | 70 |
| 1.6x per-capita GDP | 91.9 (59.2, 134) | 92.0 (60.6, 133) | 92.7 (60.7, 135) | 66 | 504 (388, 638) | 501 (389, 632) | 506 (392, 639) | 76 |
| 2.3x per-capita GDP | 120 (78.5, 174) | 120 (79.4, 172) | 121 (79.8, 174) | 70 | 660 (515, 833) | 654 (514, 823) | 662 (519, 834) | 81 |
| Woods | 41.5 (24.4, 62.7) | 42.5 (26.6, 62.9) | 42.6 (26.6, 63.3) | 51 | 219 (156, 287) | 221 (162, 286) | 222 (162, 289) | 69 |
| Ochalek | 39/0 (22.7, 59.3) | 39.9 (24.7, 59.3) | 40.0 (24.8, 59.9) | 54 | 213 (152, 280) | 213 (156, 277) | 216 (157, 282) | 71 |
| Best Buy ($100) | 29.2 (16.0, 45.3) | 30.6 (18.4, 46.1) | 30.6 (18.6, 46.2) | 47 | 155 (104, 207) | 159 (112, 208) | 159 (116, 209) | 58 |

Note: GDP = gross domestic product; No. countries = number of countries where vaccine is cost-effective at specified threshold. Values in parentheses represent equal-tailed 95% credible intervals.

### S18. Market (millions) across 2028–2050 for infant tuberculosis vaccines (societal perspective)

| | Vaccinated population where TB vaccine is cost-saving | Vaccinated population where TB vaccine is cost-effective (1x GDP per capita) |
|---|---|---|
| All countries | 1316 (1315, 1317) | 1431 (1430, 1432) |
| High-TB burden[a] | 1181 (1180, 1182) | 1232 (1231, 1232) |
| High-TB/HIV burden[a] | 815 (814, 815) | 885 (885, 886) |
| High-MDR/RR-TB burden[a] | 1019 (1018, 1020) | 1064 (1063, 1065) |
| **Income level[b]** | | |
| LIC | 182.7 (182.5, 182.8) | 195 (194, 195) |
| LMIC | 840 (839, 840) | 878 (877, 878) |
| UMIC | 294.0 (293.9, 294.0) | 359.2 (359.1, 359.2) |
| **World region** | | |
| AFR | 491.3 (490.9, 491.6) | 510.1 (509.7, 510.5) |
| AMR | 14.643 (14.641, 14.644) | 55.18 (55.17, 55.18) |
| EMR | 110.7 (110.6, 110.8) | 121.1 (121.0, 121.2) |
| EUR | 7.070 (7.069, 7.071) | 34.22 (34.219, 34.221) |
| SEAR | 401.8 (401.4, 402.0) | 406.9 (406.6, 407.2) |
| WPR | 290.9 (290.8, 291.0) | 303.7 (303.6, 303.8) |

[a] High-TB, high-TB/HIV (HIV-associated TB), and high-MDR/RR-TB (multidrug/rifampicin-resistant TB) burden countries as defined by the World Health Organization [19].

[b] LIC: Gross national income (GNI) per capita of $1,085 or less; LMIC: GNI per capita of $1,086 to $4,225; UMIC: GNI per capita of $4,256 to $13,205 (World Bank 2021).

Note: All countries include 105 low- and middle-income countries analyzed. Values in parentheses represent equal-tailed 95% credible intervals. AFR = African region; AMR = Region of the Americas; EMR = Eastern Mediterranean region; EUR = European region; GDP = gross domestic product; LIC = low-income; LMIC = lower middle-income; SEAR = Southeast Asian region; TB = tuberculosis; UMIC = upper middle-income; WPR = Western Pacific region.

### S19. Market (millions) across 2028–2050 for adolescent/adult tuberculosis vaccines (societal perspective)

| | Vaccinated population where TB vaccine is cost-saving | Vaccinated population where TB vaccine is cost-effective (1x GDP per capita) |
|---|---|---|
| All countries | 4642 (4617, 4644) | 5182 (5180, 5183) |
| High-TB burden[a] | 4008 (4006, 4009) | 4181 (4179, 4182) |
| High-TB/HIV burden[a] | 2573 (2548, 2574) | 2746 (2744, 2747) |
| High-MDR/RR-TB burden[a] | 3814 (3812, 3815) | 3868 (3867, 3869) |
| **Income level[b]** | | |
| LIC | 431 (408, 431) | 434.2 (434.0, 434.4) |
| LMIC | 2734 (2732, 2735) | 2801 (2799, 2802) |
| UMIC | 1478 (1477, 1478) | 1946.9 (1946.7, 1947.1) |
| **World region** | | |
| AFR | 1020 (996, 1020) | 1043 (1042, 1044) |
| AMR | 91.9 (91.8, 91.9) | 485.0 (485.0, 485.1) |
| EMR | 317.3 (317.1, 317.4) | 364 (363, 364) |
| EUR | 150.87 (150.86, 150.88) | 209.58 (209.56, 209.59) |
| SEAR | 1696.8 (1695.9, 1697.5) | 1715 (1714, 1716) |
| WPR | 1366 (1365, 1366) | 1366 (1365, 1366) |

[a] High-TB, high-TB/HIV (HIV-associated TB), and high-MDR/RR-TB (multidrug/rifampicin-resistant TB) burden countries as defined by the World Health Organization [19].

[b] LIC: Gross national income (GNI) per capita of $1,085 or less; LMIC: GNI per capita of $1,086 to $4,225; UMIC: GNI per capita of $4,256 to $13,205 (World Bank 2021).

Note: All countries include 105 low- and middle-income countries analyzed. Values in parentheses represent equal-tailed 95% credible intervals. AFR = African region; AMR = Region of the Americas; EMR = Eastern Mediterranean region; EUR = European region; GDP = gross domestic product; LIC = low-income; LMIC = lower middle-income; SEAR = Southeast Asian region; TB = tuberculosis; UMIC = upper middle-income; WPR = Western Pacific region.

### S20. Discounted costs, disability-adjusted life-years (DALYs) averted, and cost-effectiveness of infant tuberculosis vaccines: low-coverage scenario.

| Country grouping | Health system perspective[a] incremental cost (USD billions) | Societal perspective[b] incremental cost (USD billions) | DALYs averted (millions) | Health system cost (USD) per DALY averted | Societal cost (USD) per DALY averted |
|---|---|---|---|---|---|
| All countries | 6.76 (5.52, 9.39) | *-24.3* (*-29.0*, *-19.8*) | 16.0 (13.9, 18.5) | 426 (329, 604) | cost-saving[c] |
| High-TB burden[d] | 4.54 (3.75, 6.30) | *-24.9* (*-29.4*, *-20.6*) | 14.8 (12.6, 17.3) | 310 (239, 441) | cost-saving[c] |
| High-TB/HIV burden[d] | 3.19 (2.67, 4.30) | *-22.1* (*-26.6*, *-18.2*) | 12.9 (10.8, 15.3) | 250 (192, 349) | cost-saving[c] |
| High-MDR/RR-TB burden[d] | 4.00 (3.28, 5.59) | *-23.5* (*-27.8*, *-19.2*) | 13.0 (11.0, 15.5) | 309 (234, 447) | cost-saving[c] |
| **Income level[e]** | | | | | |
| LIC | 0.99 (0.85, 1.27) | *-0.17* (*-0.43*, 0.13) | 1.79 (1.50, 2.18) | 555 (429, 735) | cost-saving (cost-saving, 75.1) |
| LMIC | 3.48 (2.92, 4.69) | *-20.5* (*-24.6*, *-16.6*) | 13.2 (11.2, 15.6) | 266 (206, 372) | cost-saving[c] |
| UMIC | 2.29 (1.73, 3.63) | *-3.68* (*-5.53*, *-1.99*) | 0.99 (0.75, 1.33) | 2370 (1490, 3870) | cost-saving[c] |
| **World region** | | | | | |
| AFR | 2.12 (1.81, 2.76) | *-11.9* (*-15.2*, *-9.21*) | 8.29 (6.84, 10.0) | 259 (195, 351) | cost-saving[c] |
| AMR | 0.61 (0.46, 0.95) | 0.26 (0.09, 0.59) | 0.06 (0.05, 0.07) | 10900 (7680, 16900) | 4630 (1400, 10800) |
| EMR | 0.93 (0.76, 1.26) | *-0.55* (*-1.18*, *-0.02*) | 1.46 (1.02, 2.00) | 657 (428, 1010) | cost-saving[c] |
| EUR | 0.38 (0.27, 0.64) | 0.18 (0.06, 0.43) | 0.03 (0.03, 0.04) | 11200 (7550, 18900) | 5280 (1710, 13000) |
| SEAR | 1.41 (1.16, 1.92) | *-8.77* (*-11.4*, *-6.52*) | 4.87 (3.67, 6.47) | 295 (203, 449) | cost-saving[c] |
| WPR | 1.32 (1.02, 2.05) | *-3.58* (*-5.09*, *-2.33*) | 1.26 (0.92, 1.73) | 1080 (684, 1760) | cost-saving[c] |

[a] Costs from the health system perspective include vaccination costs, tuberculosis testing and treatment costs, and antiretroviral treatment costs.

[b] Costs from the societal perspective include health system perspective costs, as well as patient non-medical costs and productivity losses.

[c] Both the point estimate and the interval estimates were cost-saving.

[d] High-TB, high-TB/HIV (HIV-associated TB), and high-MDR/RR-TB (multidrug/rifampicin-resistant TB) burden countries as defined by the World Health Organization [19].

[e] LIC: Gross national income (GNI) per capita of $1,085 or less; LMIC: GNI per capita of $1,086 to $4,225; UMIC: GNI per capita of $4,256 to $13,205 (World Bank 2021).

Note: All countries include 105 low- and middle-income countries analyzed. Values in parentheses represent equal-tailed 95% credible intervals. AFR = African region; AMR = Region of the Americas; EMR = Eastern Mediterranean region; EUR = European region; LIC = low-income; LMIC = lower middle-income; SEAR = Southeast Asian region; UMIC = upper middle-income; USD = United States dollar; WPR = Western Pacific region.

### S21. Discounted costs, disability-adjusted life-years (DALYs) averted, and cost-effectiveness of adolescent/adult tuberculosis vaccines: low-coverage scenario.

| Country grouping | Health system perspective[a] incremental cost (USD billions) | Societal perspective[b] incremental cost (USD billions) | DALYs averted (millions) | Health system cost (USD) per DALY averted | Societal cost (USD) per DALY averted |
|---|---|---|---|---|---|
| All countries | 27.3 (20.3, 41.4) | *-112* (*-126*, *-94.6*) | 72.6 (66.2, 79.4) | 378 (272, 572) | cost-saving[c] |
| High-TB burden[d] | 19.5 (14.3, 30.0) | *-109* (*-123*, *-93.8*) | 66.2 (59.8, 73.1) | 296 (209, 453) | cost-saving[c] |
| High-TB/HIV burden[d] | 11.9 (8.95, 17.3) | *-101* (*-114*, *-87.8*) | 59.1 (53.2, 65.6) | 203 (145, 296) | cost-saving[c] |
| High-MDR/RR-TB burden[d] | 18.0 (12.9, 28.0) | *-102* (*-116*, *-87.7*) | 58.9 (52.7, 65.3) | 306 (213, 475) | cost-saving[c] |
| **Income level[e]** | | | | | |
| LIC | 2.88 (2.30, 3.83) | *-1.97* (*-2.84*, *-0.90*) | 7.31 (6.44, 8.22) | 395 (301, 538) | cost-saving[c] |
| LMIC | 12.9 (9.85, 18.4) | *-91.4* (*-103*, *-80.2*) | 59.9 (54.0, 66.1) | 216 (158, 311) | cost-saving[c] |
| UMIC | 11.5 (7.56, 20.4) | *-18.2* (*-25.5*, *-7.65*) | 5.37 (4.41, 6.66) | 2170 (1300, 4110) | cost-saving[c] |
| **World region** | | | | | |
| AFR | 5.34 (4.28, 7.17) | *-43.7* (*-51.5*, *-37.0*) | 30.0 (26.9, 33.3) | 179 (138, 247) | cost-saving[c] |
| AMR | 2.88 (1.95, 4.91) | *-0.10* (*-1.12*, 2.02) | 0.54 (0.48, 0.60) | 5360 (3550, 9150) | cost-saving (cost-saving, 3680) |
| EMR | 3.09 (2.43, 4.27) | *-2.07* (*-3.47*, *-0.55*) | 5.10 (4.01, 6.38) | 616 (432, 903) | cost-saving[c] |
| EUR | 1.55 (1.04, 2.66) | *-0.46* (*-1.02*, 0.63) | 0.37 (0.33, 0.42) | 4170 (2760, 7330) | cost-saving (cost-saving, 1780) |
| SEAR | 7.01 (5.24, 10.2) | *-54.6* (*-63.5*, *-45.6*) | 31.7 (27.1, 36.9) | 223 (156, 333) | cost-saving[c] |
| WPR | 7.45 (4.88, 13.1) | *-10.7* (*-14.2*, *-4.70*) | 4.85 (4.20, 5.55) | 1540 (984, 2740) | cost-saving[c] |

[a] Costs from the health system perspective include vaccination costs, tuberculosis testing and treatment costs, and antiretroviral treatment costs.
[b] Costs from the societal perspective include health system perspective costs, as well as patient non-medical costs and productivity losses.
[c] Both the point estimate and the interval estimates were cost-saving.
[d] High-TB, high-TB/HIV (HIV-associated TB), and high-MDR/RR-TB (multidrug/rifampicin-resistant TB) burden countries as defined by the World Health Organization [19].
[e] LIC: Gross national income (GNI) per capita of $1,085 or less; LMIC: GNI per capita of $1,086 to $4,225; UMIC: GNI per capita of $4,256 to $13,205 (World Bank 2021).
Note: All countries include 105 low- and middle-income countries analyzed. Values in parentheses represent equal-tailed 95% credible intervals. AFR = African region; AMR = Region of the Americas; EMR = Eastern Mediterranean region; EUR = European region; LIC = low-income; LMIC = lower middle-income; SEAR = Southeast Asian region; UMIC = upper middle-income; USD = United States dollar; WPR = Western Pacific region.

### S22. Discounted costs, disability-adjusted life-years (DALYs) averted, and cost-effectiveness of infant tuberculosis vaccines: high-coverage scenario.

| Country grouping | Health system perspective[a] incremental cost (USD billions) | Societal perspective[b] incremental cost (USD billions) | DALYs averted (millions) | Health system cost (USD) per DALY averted | Societal cost (USD) per DALY averted |
|---|---|---|---|---|---|
| All countries | 8.57 (6.99, 11.9) | *-30.6* (*-36.3*, *-24.9*) | 20.1 (17.4, 23.2) | 429 (331, 607) | cost-saving[c] |
| High-TB burden[d] | 5.76 (4.76, 7.98) | *-31.3* (*-36.8*, *-26.0*) | 18.6 (15.9, 21.7) | 312 (242, 443) | cost-saving[c] |
| High-TB/HIV burden[d] | 4.04 (3.38, 5.45) | *-27.8* (*-33.4*, *-22.9*) | 16.2 (13.6, 19.2) | 252 (194, 351) | cost-saving[c] |
| High-MDR/RR-TB burden[d] | 5.07 (4.16, 7.08) | *-29.6* (*-35.0*, *-24.2*) | 16.4 (13.8, 19.4) | 312 (237, 450) | cost-saving[c] |
| **Income level[e]** | | | | | |
| LIC | 1.25 (1.08, 1.61) | *-0.21* (*-0.53*, 0.17) | 2.25 (1.89, 2.74) | 559 (432, 738) | cost-saving (cost-saving, 78.1) |
| LMIC | 4.42 (3.70, 5.94) | *-25.7* (*-30.9*, *-20.9*) | 16.6 (14.1, 19.6) | 268 (209, 375) | cost-saving[c] |
| UMIC | 2.90 (2.19, 4.60) | *-4.65* (*-6.97*, *-2.52*) | 1.25 (0.95, 1.67) | 2380 (1490, 3870) | cost-saving[c] |
| **World region** | | | | | |
| AFR | 2.69 (2.29, 3.50) | *-14.9* (*-19.0*, *-11.6*) | 10.4 (8.61, 12.5) | 261 (197, 353) | cost-saving[c] |
| AMR | 0.77 (0.58, 1.20) | 0.32 (0.11, 0.75) | 0.07 (0.06, 0.08) | 10900 (7670, 16900) | 4600 (1380, 10700) |
| EMR | 1.18 (0.96, 1.59) | *-0.68* (*-1.47*, *-0.01*) | 1.83 (1.28, 2.51) | 663 (431, 1020) | cost-saving[c] |
| EUR | 0.48 (0.34, 0.81) | 0.22 (0.08, 0.55) | 0.04 (0.04, 0.05) | 11100 (7500, 18800) | 5210 (1660, 12800) |
| SEAR | 1.78 (1.47, 2.43) | *-11.1* (*-14.4*, *-8.24*) | 6.14 (4.64, 8.15) | 296 (204, 450) | cost-saving[c] |
| WPR | 1.68 (1.29, 2.60) | *-4.49* (*-6.40*, *-2.92*) | 1.58 (1.16, 2.17) | 1090 (690, 1770) | cost-saving[c] |

[a] Costs from the health system perspective include vaccination costs, tuberculosis testing and treatment costs, and antiretroviral treatment costs.
[b] Costs from the societal perspective include health system perspective costs, as well as patient non-medical costs and productivity losses.
[c] Both the point estimate and the interval estimates were cost-saving.
[d] High-TB, high-TB/HIV (HIV-associated TB), and high-MDR/RR-TB (multidrug/rifampicin-resistant TB) burden countries as defined by the World Health Organization [19].
[e] LIC: Gross national income (GNI) per capita of $1,085 or less; LMIC: GNI per capita of $1,086 to $4,225; UMIC: GNI per capita of $4,256 to $13,205 (World Bank 2021).
Note: All countries include 105 low- and middle-income countries analyzed. Values in parentheses represent equal-tailed 95% credible intervals. AFR = African region; AMR = Region of the Americas; EMR = Eastern Mediterranean region; EUR = European region; LIC = low-income; LMIC = lower middle-income; SEAR = Southeast Asian region; UMIC = upper middle-income; USD = United States dollar; WPR = Western Pacific region.

### S23. Discounted costs, disability-adjusted life-years (DALYs) averted, and cost-effectiveness of adolescent/adult tuberculosis vaccines: high-coverage scenario.

| Country grouping | Health system perspective[a] incremental cost (USD billions) | Societal perspective[b] incremental cost (USD billions) | DALYs averted (millions) | Health system cost (USD) per DALY averted | Societal cost (USD) per DALY averted |
|---|---|---|---|---|---|
| All countries | 45.2 (33.4, 69.0) | *-178* (*-201*, *-150*) | 116 (107, 127) | 390 (280, 592) | cost-saving[c] |
| High-TB burden[d] | 32.4 (23.7, 49.8) | *-173* (*-195*, *-149*) | 106 (96.5, 117) | 307 (218, 469) | cost-saving[c] |
| High-TB/HIV burden[d] | 19.6 (14.7, 28.6) | *-161* (*-180*, *-141*) | 94.5 (85.5, 104) | 208 (150, 304) | cost-saving[c] |
| High-MDR/RR-TB burden[d] | 30.0 (21.5, 46.9) | *-163* (*-184*, *-139*) | 94.3 (85.1, 104) | 319 (221, 497) | cost-saving[c] |
| **Income level[e]** | | | | | |
| LIC | 4.59 (3.67, 6.13) | *-3.12* (*-4.49*, *-1.44*) | 11.7 (10.3, 13.1) | 396 (303, 538) | cost-saving[c] |
| LMIC | 21.3 (16.2, 30.2) | *-145* (*-162*, *-128*) | 95.8 (86.8, 105) | 222 (163, 322) | cost-saving[c] |
| UMIC | 19.4 (12.7, 34.4) | *-29.7* (*-41.8*, *-12.1*) | 8.90 (7.35, 11.0) | 2200 (1330, 4170) | cost-saving[c] |
| **World region** | | | | | |
| AFR | 8.45 (6.75, 11.3) | *-69.0* (*-80.7*, *-59.2*) | 47.5 (42.9, 52.1) | 179 (138, 246) | cost-saving[c] |
| AMR | 4.81 (3.25, 8.22) | *-0.22* (*-1.92*, 3.33) | 0.91 (0.82, 1.02) | 5280 (3510, 9010) | cost-saving (cost-saving, 3390) |
| EMR | 5.04 (3.95, 6.95) | *-3.16* (*-5.37*, *-0.76*) | 8.07 (6.39, 10.0) | 632 (444, 919) | cost-saving[c] |
| EUR | 2.60 (1.75, 4.47) | *-0.82* (*-1.78*, 1.02) | 0.63 (0.57, 0.71) | 4120 (2710, 7240) | cost-saving (cost-saving, 1690) |
| SEAR | 11.8 (8.79, 17.1) | *-87.4* (*-101*, *-73.5*) | 51.4 (44.2, 59.3) | 231 (162, 344) | cost-saving[c] |
| WPR | 12.6 (8.23, 22.1) | *-16.9* (*-22.7*, *-6.73*) | 7.87 (6.86, 9.04) | 1610 (1020, 2860) | cost-saving[c] |

[a] Costs from the health system perspective include vaccination costs, tuberculosis testing and treatment costs, and antiretroviral treatment costs.
[b] Costs from the societal perspective include health system perspective costs, as well as patient non-medical costs and productivity losses.
[c] Both the point estimate and the interval estimates were cost-saving.
[d] High-TB, high-TB/HIV (HIV-associated TB), and high-MDR/RR-TB (multidrug/rifampicin-resistant TB) burden countries as defined by the World Health Organization [19].
[e] LIC: Gross national income (GNI) per capita of $1,085 or less; LMIC: GNI per capita of $1,086 to $4,225; UMIC: GNI per capita of $4,256 to $13,205 (World Bank 2021).
Note: All countries include 105 low- and middle-income countries analyzed. Values in parentheses represent equal-tailed 95% credible intervals. AFR = African region; AMR = Region of the Americas; EMR = Eastern Mediterranean region; EUR = European region; LIC = low-income; LMIC = lower middle-income; SEAR = Southeast Asian region; UMIC = upper middle-income; USD = United States dollar; WPR = Western Pacific region.

### S24. Discounted costs, disability-adjusted life-years (DALYs) averted, and cost-effectiveness of infant tuberculosis vaccines: accelerated scale-up scenario.

| Country grouping | Health system perspective[a] incremental cost (USD billions) | Societal perspective[b] incremental cost (USD billions) | DALYs averted (millions) | Health system cost (USD) per DALY averted | Societal cost (USD) per DALY averted |
|---|---|---|---|---|---|
| All countries | *-1.15* (*-3.76*, 3.51) | *-5480* (*-5490*, *-5470*) | 1480 (1480, 1490) | cost-saving (cost-saving, 2.38) | cost-saving[c] |
| High-TB burden[d] | *-2.85* (*-4.73*, 0.16) | *-4190* (*-4200*, *-4170*) | 1130 (1130, 1140) | cost-saving (cost-saving, 0.14) | cost-saving[c] |
| High-TB/HIV burden[d] | *-6.05* (*-7.47*, *-3.93*) | *-1890* (*-1900*, *-1870*) | 770 (764, 778) | cost-saving[c] | cost-saving[c] |
| High-MDR/RR-TB burden[d] | *-1.89* (*-3.61*, 0.93) | *-4230* (*-4250*, *-4220*) | 1100 (1090, 1100) | cost-saving (cost-saving, 0.85) | cost-saving[c] |
| **Income level[e]** | | | | | |
| LIC | 0.003 (*-0.34*, 0.60) | *-94.8* (*-95.8*, *-94.0*) | 136 (134, 137) | 0.02 (cost-saving, 4.39) | cost-saving[c] |
| LMIC | 0.71 (*-0.75*, 3.03) | *-1450* (*-1460*, *-1440*) | 806 (800, 814) | 0.88 (cost-saving, 3.77) | cost-saving[c] |
| UMIC | *-1.86* (*-2.91*, 0.20) | *-3940* (*-3940*, *-3930*) | 540 (538, 541) | cost-saving (cost-saving, 0.38) | cost-saving[c] |
| **World region** | | | | | |
| AFR | *-6.69* (*-7.62*, *-5.43*) | *-525* (*-535*, *-516*) | 314 (310, 319) | cost-saving[c] | cost-saving[c] |
| AMR | 0.96 (0.71, 1.52) | *-685* (*-685*, *-684*) | 105.8 (105.7, 105.8) | 9.11 (6.69, 14.3) | cost-saving[c] |
| EMR | 1.42 (1.10, 1.98) | *-203* (*-204*, *-202*) | 128 (127, 129) | 11.1 (8.61, 15.4) | cost-saving[c] |
| EUR | 0.46 (0.27, 0.88) | *-519.9* (*-520.1*, *-519.5*) | 90.1 (90.1, 90.2) | 5.14 (3.02, 9.74) | cost-saving[c] |
| SEAR | 1.28 (0.50, 2.41) | *-879* (*-889*, *-871*) | 469 (465, 475) | 2.73 (1.06, 5.13) | cost-saving[c] |
| WPR | 1.41 (0.90, 2.51) | *-2667* (*-2670*, *-2665*) | 374 (373, 375) | 3.76 (2.40, 6.69) | cost-saving[c] |

[a] Costs from the health system perspective include vaccination costs, tuberculosis testing and treatment costs, and antiretroviral treatment costs.
[b] Costs from the societal perspective include health system perspective costs, as well as patient non-medical costs and productivity losses.
[c] Both the point estimate and the interval estimates were cost-saving.
[d] High-TB, high-TB/HIV (HIV-associated TB), and high-MDR/RR-TB (multidrug/rifampicin-resistant TB) burden countries as defined by the World Health Organization [19].
[e] LIC: Gross national income (GNI) per capita of $1,085 or less; LMIC: GNI per capita of $1,086 to $4,225; UMIC: GNI per capita of $4,256 to $13,205 (World Bank 2021).
Note: All countries include 105 low- and middle-income countries analyzed. Values in parentheses represent equal-tailed 95% credible intervals. AFR = African region; AMR = Region of the Americas; EMR = Eastern Mediterranean region; EUR = European region; LIC = low-income; LMIC = lower middle-income; SEAR = Southeast Asian region; UMIC = upper middle-income; USD = United States dollar; WPR = Western Pacific region.

**S25. Discounted costs, disability-adjusted life-years (DALYs) averted, and cost-effectiveness of adolescent/adult tuberculosis vaccines: accelerated scale-up scenario.**

| Country grouping | Health system perspective[a] incremental cost (USD billions) | Societal perspective[b] incremental cost (USD billions) | DALYs averted (millions) | Health system cost (USD) per DALY averted | Societal cost (USD) per DALY averted |
|---|---|---|---|---|---|
| All countries | 28.7 (17.4, 51.6) | *-5670* (*-5700*, *-5640*) | 1600 (1580, 1610) | 18.0 (10.9, 32.1) | cost-saving[c] |
| High-TB burden[d] | 19.0 (10.5, 35.5) | *-4370* (*-4400*, *-4330*) | 1240 (1220, 1250) | 15.4 (8.51, 28.6) | cost-saving[c] |
| High-TB/HIV burden[d] | 6.45 (1.34, 15.3) | *-2050* (*-2080*, *-2020*) | 863 (848, 878) | 7.48 (1.55, 17.6) | cost-saving[c] |
| High-MDR/RR-TB burden[d] | 18.6 (10.4, 34.8) | *-4400* (*-4430*, *-4370*) | 1190 (1180, 1210) | 15.6 (8.72, 29.3) | cost-saving[c] |
| **Income level[e]** | | | | | |
| LIC | 2.49 (1.57, 3.99) | *-100* (*-101*, *-97.7*) | 147 (145, 149) | 17.0 (10.7, 27.3) | cost-saving[c] |
| LMIC | 14.2 (8.88, 23.3) | *-1600* (*-1630*, *-1580*) | 903 (888, 917) | 15.8 (9.83, 25.9) | cost-saving[c] |
| UMIC | 12.0 (5.93, 25.5) | *-3970* (*-3980*, *-3950*) | 548 (546, 551) | 21.9 (10.8, 46.7) | cost-saving[c] |
| **World region** | | | | | |
| AFR | *-2.45* (*-4.23*, 0.36) | *-582* (*-600*, *-567*) | 353 (347, 360) | cost-saving (cost-saving, 1.01) | cost-saving[c] |
| AMR | 4.38 (2.94, 7.53) | *-688* (*-689*, *-684*) | 106.9 (106.7, 107.1) | 41.0 (27.4, 70.5) | cost-saving[c] |
| EMR | 4.47 (3.44, 6.34) | *-207* (*-209*, *-204*) | 135 (133, 137) | 33.2 (25.5, 46.5) | cost-saving[c] |
| EUR | 2.23 (1.41, 3.98) | *-523* (*-524*, *-521*) | 91.1 (90.9, 91.2) | 24.5 (15.5, 43.6) | cost-saving[c] |
| SEAR | 9.54 (6.24, 14.9) | *-988* (*-1010*, *-966*) | 530 (518, 543) | 18.0 (11.7, 28.4) | cost-saving[c] |
| WPR | 10.6 (6.60, 19.0) | *-2680* (*-2690*, *-2680*) | 381 (380, 383) | 27.7 (17.3, 50.0) | cost-saving[c] |

[a] Costs from the health system perspective include vaccination costs, tuberculosis testing and treatment costs, and antiretroviral treatment costs.

[b] Costs from the societal perspective include health system perspective costs, as well as patient non-medical costs and productivity losses.

[c] Both the point estimate and the interval estimates were cost-saving.

[d] High-TB, high-TB/HIV (HIV-associated TB), and high-MDR/RR-TB (multidrug/rifampicin-resistant TB) burden countries as defined by the World Health Organization [19].

[e] LIC: Gross national income (GNI) per capita of $1,085 or less; LMIC: GNI per capita of $1,086 to $4,225; UMIC: GNI per capita of $4,256 to $13,205 (World Bank 2021).

Note: All countries include 105 low- and middle-income countries analyzed. Values in parentheses represent equal-tailed 95% credible intervals. AFR = African region; AMR = Region of the Americas; EMR = Eastern Mediterranean region; EUR = European region; LIC = low-income; LMIC = lower middle-income; SEAR = Southeast Asian region; UMIC = upper middle-income; USD = United States dollar; WPR = Western Pacific region.

**S26. Percentage of population that live in countries where vaccination was cost-effective from the health system perspective compared to percentage of gross domestic product per capita thresholds, comparing alternative scale-up scenarios.**

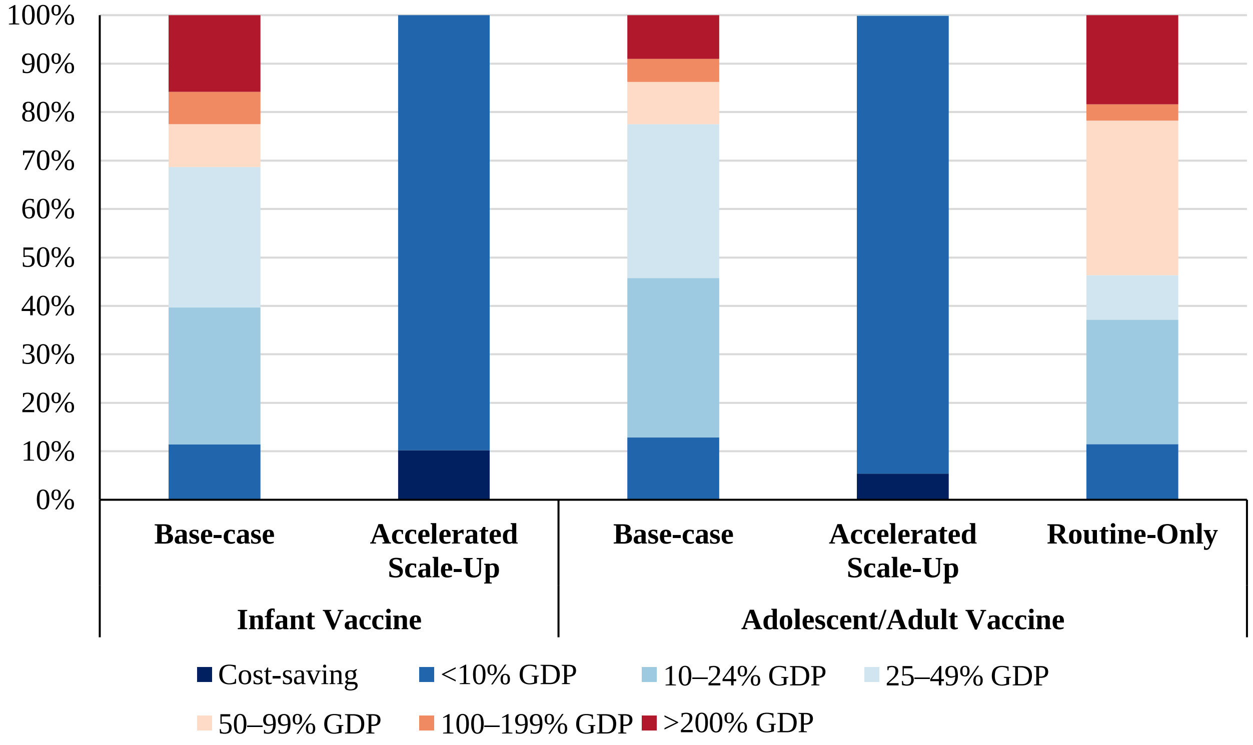


Note: Countries include 105 low- and middle-income countries analyzed. Population includes vaccinated individuals 2028–2050. Gross domestic product per capita estimates from 2020. GDP = gross domestic product per capita.

**S27. Percentage of countries where vaccination was cost-effective from the health system perspective compared to percentage of gross domestic product per capita thresholds, comparing alternative scale-up scenarios.**

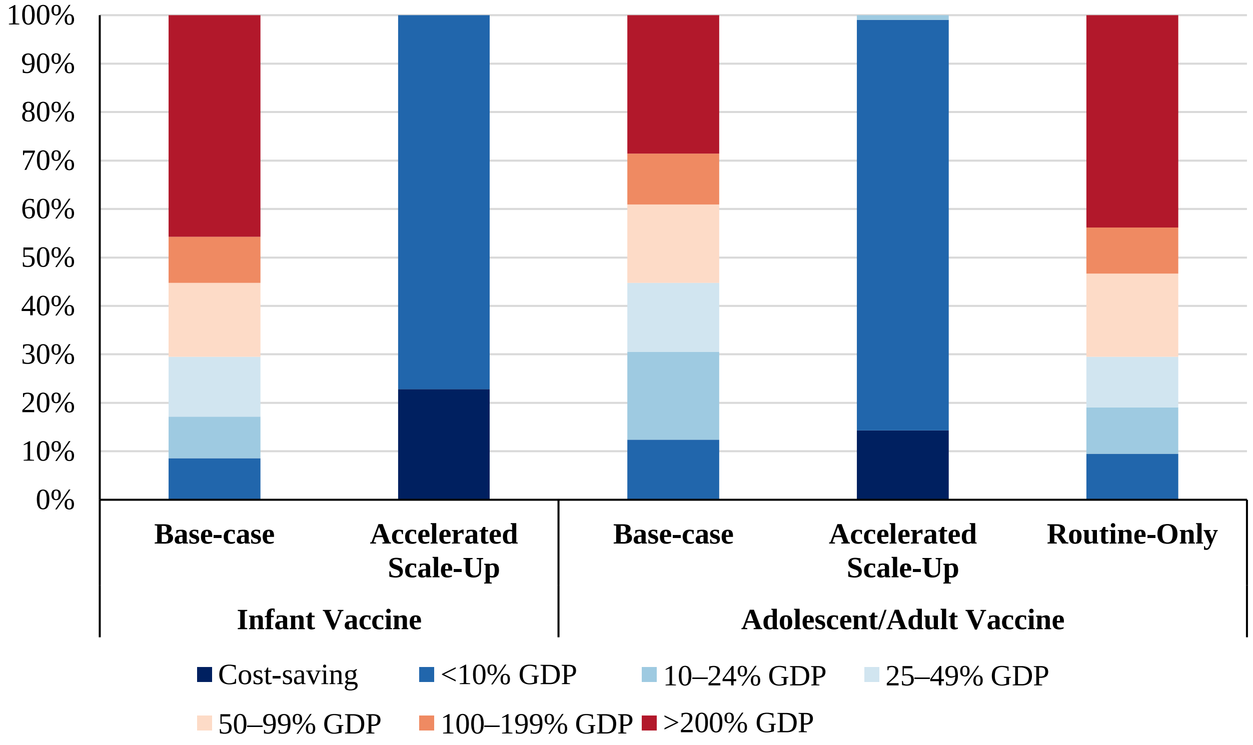


Note: Countries include 105 low- and middle-income countries analyzed. Gross domestic product per capita estimates from 2020. GDP = gross domestic product per capita.

### S28. Discounted costs, disability-adjusted life-years (DALYs) averted, and cost-effectiveness of adolescent tuberculosis vaccines: routine-only scale-up scenario.

| Country grouping | Health system perspective[a] incremental cost (USD billions) | Societal perspective[b] incremental cost (USD billions) | DALYs averted (millions) | Health system cost (USD) per DALY averted | Societal cost (USD) per DALY averted |
|---|---|---|---|---|---|
| All countries | 9.15 (7.39, 12.4) | -31.8 (-38.5, -25.3) | 21.2 (18.3, 24.8) | 435 (326, 609) | cost-saving[c] |
| High-TB burden[d] | 6.38 (5.10, 8.63) | -32.5 (-39.0, -26.1) | 19.7 (16.8, 23.2) | 327 (239, 465) | cost-saving[c] |
| High-TB/HIV burden[d] | 4.40 (3.59, 5.85) | -30.2 (-36.8, -24.9) | 17.6 (14.8, 21.3) | 252 (187, 349) | cost-saving[c] |
| High-MDR/RR-TB burden[d] | 5.62 (4.45, 7.74) | -30.8 (-37.4, -24.6) | 17.4 (14.6, 21.0) | 326 (233, 472) | cost-saving[c] |
| **Income level[e]** | | | | | |
| LIC | 1.33 (1.12, 1.68) | -0.14 (-0.49, 0.25) | 2.21 (1.88, 2.63) | 605 (466, 792) | cost-saving (cost-saving, 122) |
| LMIC | 4.75 (3.89, 6.21) | -28.0 (-34.4, -22.9) | 17.8 (15.0, 21.4) | 269 (200, 366) | cost-saving[c] |
| UMIC | 3.07 (2.24, 4.89) | -3.70 (-6.12, -1.36) | 1.18 (0.88, 1.60) | 2680 (1610, 4500) | cost-saving[c] |
| **World region** | | | | | |
| AFR | 2.73 (2.31, 3.45) | -14.4 (-19.0, -10.8) | 10.2 (8.41, 12.6) | 271 (200, 366) | cost-saving[c] |
| AMR | 0.83 (0.62, 1.30) | 0.39 (0.15, 0.87) | 0.07 (0.06, 0.09) | 11400 (7880, 17800) | 5330 (1860, 11700) |
| EMR | 1.23 (1.02, 1.59) | -0.43 (-1.02, 0.14) | 1.66 (1.21, 2.22) | 760 (520, 1110) | cost-saving (cost-saving, 92.5) |
| EUR | 0.44 (0.33, 0.69) | 0.19 (0.07, 0.44) | 0.05 (0.04, 0.05) | 9390 (6720, 15000) | 4090 (1480, 9760) |
| SEAR | 2.06 (1.65, 2.73) | -14.3 (-18.4, -10.8) | 7.77 (5.93, 10.2) | 270 (181, 399) | cost-saving[c] |
| WPR | 1.86 (1.36, 2.93) | -3.27 (-4.50, -2.00) | 1.43 (1.13, 1.79) | 1320 (858, 2120) | cost-saving[c] |

[a] Costs from the health system perspective include vaccination costs, tuberculosis testing and treatment costs, and antiretroviral treatment costs.
[b] Costs from the societal perspective include health system perspective costs, as well as patient non-medical costs and productivity losses.
[c] Both the point estimate and the interval estimates were cost-saving.
[d] High-TB, high-TB/HIV (HIV-associated TB), and high-MDR/RR-TB (multidrug/rifampicin-resistant TB) burden countries as defined by the World Health Organization [19].
[e] LIC: Gross national income (GNI) per capita of $1,085 or less; LMIC: GNI per capita of $1,086 to $4,225; UMIC: GNI per capita of $4,256 to $13,205 (World Bank 2021).
Note: All countries include 105 low- and middle-income countries analyzed. Values in parentheses represent equal-tailed 95% credible intervals. AFR = African region; AMR = Region of the Americas; EMR = Eastern Mediterranean region; EUR = European region; LIC = low-income; LMIC = lower middle-income; SEAR = Southeast Asian region; UMIC = upper middle-income; USD = United States dollar; WPR = Western Pacific region.

**S29. Undiscounted vaccination costs (billions) of infant tuberculosis vaccines across 2028–2050 by vaccine price and dose scenario.**

| Country grouping | Base-case | Half-price | Double-price | High-middle-tier-price | Two-dose |
|---|---|---|---|---|---|
| Global | 11.8 (9.59–16.9) | 7.59 (5.40–12.7) | 20.2 (18.0–25.2) | 16.9 (14.7–22.0) | 23.3 (18.9–33.4) |
| High-TB burden[a] | 7.99 (6.65–11.2) | 4.79 (3.51–7.85) | 13.3 (12.0–16.4) | 10.75 (9.47–13.8) | 15.1 (12.5–21.2) |
| High-TB/HIV burden[a] | 5.71 (4.80–7.81) | 3.55 (2.64–5.65) | 10.0 (9.12–12.1) | 6.73 (5.82–8.83) | 11.3 (9.46–15.5) |
| High-MDR/RR-TB burden[a] | 7.06 (5.85–10.0) | 4.45 (3.24–7.43) | 12.3 (11.1–15.3) | 10.3 (9.07–13.3) | 14.0 (11.5–19.9) |
| **Income level[b]** | | | | | |
| LIC | 1.84 (1.57–2.41) | 1.07 (0.82–1.61) | 3.09 (2.85–3.63) | 1.75 (1.50–2.28) | 3.45 (2.95–4.52) |
| LMIC | 5.68 (4.80–7.65) | 3.60 (2.70–5.60) | 10.1 (9.22–12.1) | 6.52 (5.61–8.52) | 11.4 (9.59–15.4) |
| UMIC | 4.26 (3.22–6.8) | 2.92 (1.88–5.46) | 6.95 (5.91–9.49) | 8.65 (7.61–11.2) | 8.43 (6.35–13.5) |
| **World region** | | | | | |
| AFR | 3.73 (3.16–4.99) | 2.32 (1.75–3.58) | 6.56 (5.99–7.82) | 3.87 (3.30–5.13) | 7.37 (6.23–9.89) |
| AMR | 0.99 (0.75–1.56) | 0.68 (0.44–1.25) | 1.61 (1.36–2.18) | 1.93 (1.69–2.50) | 1.96 (1.47–3.10) |
| EMR | 1.64 (1.35–2.25) | 1.06 (0.77–1.67) | 2.80 (2.51–3.41) | 2.40 (2.11–3.01) | 3.24 (2.66–4.46) |
| EUR | 0.63 (0.44–1.07) | 0.46 (0.27–0.90) | 0.97 (0.78–1.42) | 1.09 (0.90–1.53) | 1.25 (0.87–2.13) |
| SEAR | 2.60 (2.18–3.54) | 1.62 (1.20–2.56) | 4.56 (4.15–5.51) | 2.65 (2.23–3.59) | 5.13 (4.29–7.01) |
| WPR | 2.19 (1.71–3.44) | 1.46 (0.98–2.71) | 3.66 (3.18–4.91) | 4.97 (4.49–6.22) | 4.34 (3.37–6.84) |

[a] High-TB, high-TB/HIV (HIV-associated TB), and high-MDR/RR-TB (multidrug/rifampicin-resistant TB) burden countries as defined by the World Health Organization [19].

[b] LIC: Gross national income (GNI) per capita of $1,085 or less; LMIC: GNI per capita of $1,086 to $4,225; UMIC: GNI per capita of $4,256 to $13,205 (World Bank 2021).

Note: All countries include 105 low- and middle-income countries analyzed. Values in parentheses represent equal-tailed 95% credible intervals. AFR = African region; AMR = Region of the Americas; EMR = Eastern Mediterranean region; EUR = European region; LIC = low-income; LMIC = lower middle-income; SEAR = Southeast Asian region; TB = tuberculosis; UMIC = upper middle-income; WPR = Western Pacific region.

### S30. Undiscounted vaccination costs (billions) of adolescent/adult tuberculosis vaccines across 2028–2050 by vaccine price and dose scenario.

| Country grouping | Base-case | Half-price | Double-price | High-middle-tier-price | Two-dose |
|---|---|---|---|---|---|
| Global | 50.5 (38.1–75.9) | 36.4 (24.0–61.8) | 78.8 (66.4–104.2) | 72.1 (59.7–97.5) | 100 (75.3–151) |
| High-TB burden[a] | 36.6 (27.7–54.7) | 24.9 (16.5–42.0) | 53.9 (45.5–71.0) | 48.9 (40.5–66.0) | 68.5 (51.7–103) |
| High-TB/HIV burden[a] | 23.5 (18.4–33.3) | 16.8 (11.7–26.6) | 37.0 (31.9–46.7) | 27.5 (22.4–37.2) | 46.6 (36.4–66.1) |
| High-MDR/RR-TB burden[a] | 34.1 (25.8–51.4) | 24.6 (16.2–41.9) | 53.2 (44.8–70.5) | 48.7 (40.3–66.0) | 67.7 (50.9–102) |
| **Income level[b]** | | | | | |
| LIC | 5.60 (4.50–7.46) | 3.81 (2.75–5.61) | 8.57 (7.51–10.4) | 5.40 (4.33–7.20) | 10.7 (8.55–14.3) |
| LMIC | 23.1 (18.6–31.1) | 16.5 (12.1–24.6) | 36.7 (32.2–44.8) | 25.6 (21.1–33.7) | 46.1 (37.1–62.2) |
| UMIC | 21.9 (15.0–37.4) | 16.0 (9.20–31.6) | 33.5 (26.7–49.0) | 41.1 (34.3–56.6) | 43.3 (29.7–74.4) |
| **World region** | | | | | |
| AFR | 9.96 (7.98–13.5) | 7.07 (5.09–10.6) | 15.7 (13.8–19.3) | 10.4 (8.38–13.9) | 19.7 (15.7–26.8) |
| AMR | 4.78 (3.29–8.17) | 3.51 (2.02–6.90) | 7.33 (5.84–10.7) | 8.53 (7.04–11.9) | 9.48 (6.49–16.3) |
| EMR | 5.66 (4.49–7.86) | 4.04 (2.86–6.23) | 8.91 (7.74–11.1) | 8.01 (6.84–10.2) | 11.2 (8.86–15.6) |
| EUR | 2.82 (1.98–4.72) | 2.06 (1.23–3.96) | 4.34 (3.50–6.23) | 4.79 (3.96–6.69) | 5.59 (3.92–9.38) |
| SEAR | 14.6 (11.7–20.0) | 10.4 (7.46–15.8) | 23.0 (20.1–28.4) | 14.9 (12.0–20.3) | 28.9 (23.0–39.7) |
| WPR | 12.7 (8.73–21.7) | 9.30 (5.35–18.3) | 19.4 (15.5–28.5) | 25.4 (21.5–34.5) | 25.1 (17.2–43.2) |

[a] High-TB, high-TB/HIV (HIV-associated TB), and high-MDR/RR-TB (multidrug/rifampicin-resistant TB) burden countries as defined by the World Health Organization [19].

[b] LIC: Gross national income (GNI) per capita of $1,085 or less; LMIC: GNI per capita of $1,086 to $4,225; UMIC: GNI per capita of $4,256 to $13,205 (World Bank 2021).

Note: All countries include 105 low- and middle-income countries analyzed. Values in parentheses represent equal-tailed 95% credible intervals. AFR = African region; AMR = Region of the Americas; EMR = Eastern Mediterranean region; EUR = European region; LIC = low-income; LMIC = lower middle-income; SEAR = Southeast Asian region; TB = tuberculosis; UMIC = upper middle-income; WPR = Western Pacific region.

**S31. Discounted costs, disability-adjusted life-years (DALYs) averted, and cost-effectiveness of infant tuberculosis vaccines: half-price scenario.**

| Country grouping | Health system perspective[a] incremental cost (USD billions) | Societal perspective[b] incremental cost (USD billions) | DALYs averted (millions) | Health system cost (USD) per DALY averted | Societal cost (USD) per DALY averted |
|---|---|---|---|---|---|
| All countries | 4.81 (3.40, 7.79) | *-30.3* (*-35.5*, *-25.2*) | 18.0 (15.6, 20.8) | 269 (183, 438) | cost-saving[c] |
| High-TB burden[d] | 3.11 (2.22, 5.10) | *-30.1* (*-35.1*, *-25.3*) | 16.6 (14.2, 19.5) | 188 (128, 313) | cost-saving[c] |
| High-TB/HIV burden[d] | 2.15 (1.56, 3.41) | *-26.4* (*-31.4*, *-22.0*) | 14.5 (12.2, 17.3) | 149 (102, 240) | cost-saving[c] |
| High-MDR/RR-TB burden[d] | 2.74 (1.93, 4.54) | *-28.3* (*-33.2*, *-23.5*) | 14.7 (12.4, 17.4) | 188 (126, 323) | cost-saving[c] |
| **Income level[e]** | | | | | |
| LIC | 0.68 (0.52, 1.00) | *-0.63* (*-0.92*, *-0.29*) | 2.02 (1.70, 2.46) | 337 (242, 501) | cost-saving[c] |
| LMIC | 2.36 (1.72, 3.72) | *-24.7* (*-29.3*, *-20.4*) | 14.9 (12.6, 17.6) | 160 (110, 258) | cost-saving[c] |
| UMIC | 1.78 (1.14, 3.30) | *-4.97* (*-7.06*, *-3.07*) | 1.12 (0.85, 1.50) | 1630 (902, 3100) | cost-saving[c] |
| **World region** | | | | | |
| AFR | 1.45 (1.10, 2.18) | *-14.3* (*-18.0*, *-11.3*) | 9.34 (7.72, 11.3) | 157 (109, 243) | cost-saving[c] |
| AMR | 0.47 (0.31, 0.86) | 0.08 (*-0.11*, 0.46) | 0.06 (0.05, 0.08) | 7490 (4550, 13500) | 1240 (cost-saving, 7180) |
| EMR | 0.66 (0.47, 1.03) | *-1.01* (*-1.71*, *-0.40*) | 1.64 (1.15, 2.25) | 415 (247, 713) | cost-saving[c] |
| EUR | 0.31 (0.18, 0.60) | 0.08 (*-0.05*, 0.37) | 0.04 (0.03, 0.04) | 8080 (4610, 15600) | 2210 (cost-saving, 9580) |
| SEAR | 0.93 (0.66, 1.52) | *-10.6* (*-13.6*, *-8.03*) | 5.50 (4.15, 7.31) | 174 (107, 302) | cost-saving[c] |
| WPR | 0.98 (0.64, 1.81) | *-4.55* (*-6.26*, *-3.14*) | 1.42 (1.04, 1.95) | 708 (391, 1330) | cost-saving[c] |

[a] Costs from the health system perspective include vaccination costs, tuberculosis testing and treatment costs, and antiretroviral treatment costs.
[b] Costs from the societal perspective include health system perspective costs, as well as patient non-medical costs and productivity losses.
[c] Both the point estimate and the interval estimates were cost-saving.
[d] High-TB, high-TB/HIV (HIV-associated TB), and high-MDR/RR-TB (multidrug/rifampicin-resistant TB) burden countries as defined by the World Health Organization [19].
[e] LIC: Gross national income (GNI) per capita of $1,085 or less; LMIC: GNI per capita of $1,086 to $4,225; UMIC: GNI per capita of $4,256 to $13,205 (World Bank 2021).
Note: All countries include 105 low- and middle-income countries analyzed. Values in parentheses represent equal-tailed 95% credible intervals. AFR = African region; AMR = Region of the Americas; EMR = Eastern Mediterranean region; EUR = European region; LIC = low-income; LMIC = lower middle-income; SEAR = Southeast Asian region; UMIC = upper middle-income; USD = United States dollar; WPR = Western Pacific region.

**S32. Discounted costs, disability-adjusted life-years (DALYs) averted, and cost-effectiveness of adolescent/adult tuberculosis vaccines: half-price scenario.**

| Country grouping | Health system perspective[a] incremental cost (USD billions) | Societal perspective[b] incremental cost (USD billions) | DALYs averted (millions) | Health system cost (USD) per DALY averted | Societal cost (USD) per DALY averted |
|---|---|---|---|---|---|
| All countries | 25.0 (15.6, 43.9) | *-156* (*-176*, *-135*) | 94.8 (86.9, 103) | 264 (161, 462) | cost-saving[c] |
| High-TB burden[d] | 17.7 (10.7, 31.6) | *-150* (*-168*, *-131*) | 86.5 (78.5, 95.1) | 205 (123, 361) | cost-saving[c] |
| High-TB/HIV burden[d] | 10.5 (6.53, 17.6) | *-137* (*-153*, *-120*) | 77.1 (69.6, 85.3) | 136 (82, 231) | cost-saving[c] |
| High-MDR/RR-TB burden[d] | 16.3 (9.48, 29.8) | *-141* (*-158*, *-122*) | 76.9 (69.2, 85.3) | 212 (123, 383) | cost-saving[c] |
| **Income level[e]** | | | | | |
| LIC | 2.58 (1.83, 3.83) | *-3.72* (*-4.85*, *-2.35*) | 9.53 (8.40, 10.7) | 272 (188, 408) | cost-saving[c] |
| LMIC | 11.3 (7.24, 18.5) | *-124* (*-139*, *-110*) | 78.2 (70.8, 86.3) | 145 (89.9, 244) | cost-saving[c] |
| UMIC | 11.1 (5.78, 23.1) | *-28.3* (*-38.1*, *-14.3*) | 7.15 (5.89, 8.83) | 1570 (785, 3490) | cost-saving[c] |
| **World region** | | | | | |
| AFR | 4.71 (3.33, 7.07) | *-58.8* (*-68.6*, *-50.5*) | 38.9 (35.0, 42.8) | 121 (83.8, 186) | cost-saving[c] |
| AMR | 2.79 (1.55, 5.52) | *-1.22* (*-2.58*, 1.62) | 0.73 (0.65, 0.81) | 3850 (2120, 7680) | cost-saving (cost-saving, 2180) |
| EMR | 2.80 (1.92, 4.34) | *-3.91* (*-5.72*, *-1.97*) | 6.61 (5.22, 8.24) | 429 (271, 702) | cost-saving[c] |
| EUR | 1.46 (0.78, 2.94) | *-1.26* (*-2.02*, 0.20) | 0.50 (0.45, 0.56) | 2900 (1530, 5960) | cost-saving (cost-saving, 424) |
| SEAR | 6.12 (3.75, 10.4) | *-74.6* (*-85.9*, *-63.1*) | 41.7 (35.8, 48.3) | 148 (84.4, 255) | cost-saving[c] |
| WPR | 7.11 (3.65, 14.7) | *-16.7* (*-21.4*, *-8.69*) | 6.37 (5.55, 7.31) | 1120 (568, 2330) | cost-saving[c] |

[a] Costs from the health system perspective include vaccination costs, tuberculosis testing and treatment costs, and antiretroviral treatment costs.
[b] Costs from the societal perspective include health system perspective costs, as well as patient non-medical costs and productivity losses.
[c] Both the point estimate and the interval estimates were cost-saving.
[d] High-TB, high-TB/HIV (HIV-associated TB), and high-MDR/RR-TB (multidrug/rifampicin-resistant TB) burden countries as defined by the World Health Organization [19].
[e] LIC: Gross national income (GNI) per capita of $1,085 or less; LMIC: GNI per capita of $1,086 to $4,225; UMIC: GNI per capita of $4,256 to $13,205 (World Bank 2021).
Note: All countries include 105 low- and middle-income countries analyzed. Values in parentheses represent equal-tailed 95% credible intervals. AFR = African region; AMR = Region of the Americas; EMR = Eastern Mediterranean region; EUR = European region; LIC = low-income; LMIC = lower middle-income; SEAR = Southeast Asian region; UMIC = upper middle-income; USD = United States dollar; WPR = Western Pacific region.

### S33. Discounted costs, disability-adjusted life-years (DALYs) averted, and cost-effectiveness of infant tuberculosis vaccines: double-price scenario.

| Country grouping | Health system perspective[a] incremental cost (USD billions) | Societal perspective[b] incremental cost (USD billions) | DALYs averted (millions) | Health system cost (USD) per DALY averted | Societal cost (USD) per DALY averted |
|---|---|---|---|---|---|
| All countries | 13.4 (12.0, 16.3) | *-21.7* (*-26.9*, *-16.6*) | 18.0 (15.6, 20.8) | 746 (619, 934) | cost-saving[c] |
| High-TB burden[d] | 9.23 (8.33, 11.2) | *-24.0* (*-29.0*, *-19.2*) | 16.6 (14.2, 19.5) | 558 (459, 696) | cost-saving[c] |
| High-TB/HIV burden[d] | 6.55 (5.96, 7.81) | *-22.0* (*-27.0*, *-17.6*) | 14.5 (12.2, 17.3) | 454 (369, 576) | cost-saving[c] |
| High-MDR/RR-TB burden[d] | 8.11 (7.3, 9.91) | *-22.9* (*-27.8*, *-18.1*) | 14.7 (12.4, 17.4) | 556 (449, 705) | cost-saving[c] |
| **Income level[e]** | | | | | |
| LIC | 2.00 (1.85, 2.32) | 0.69 (0.41, 1.03) | 2.02 (1.70, 2.46) | 999 (801, 1236) | 350 (173, 573) |
| LMIC | 7.13 (6.49, 8.49) | *-19.9* (*-24.6*, *-15.6*) | 14.9 (12.6, 17.6) | 483 (390, 604) | cost-saving[c] |
| UMIC | 4.23 (3.59, 5.76) | *-2.52* (*-4.60*, *-0.61*) | 1.12 (0.85, 1.50) | 3870 (2670, 5620) | cost-saving[c] |
| **World region** | | | | | |
| AFR | 4.30 (3.95, 5.03) | *-11.5* (*-15.2*, *-8.49*) | 9.34 (7.72, 11.3) | 465 (367, 593) | cost-saving[c] |
| AMR | 1.12 (0.95, 1.50) | 0.72 (0.53, 1.10) | 0.06 (0.05, 0.08) | 17700 (13600, 24200) | 11418 (7313, 17880) |
| EMR | 1.84 (1.64, 2.21) | 0.17 (*-0.54*, 0.77) | 1.64 (1.15, 2.25) | 1150 (786, 1670) | 138 (cost-saving, 660) |
| EUR | 0.67 (0.54, 0.96) | 0.44 (0.31, 0.72) | 0.04 (0.03, 0.04) | 17200 (13200, 25300) | 11361 (7389, 19458) |
| SEAR | 2.91 (2.63, 3.49) | *-8.58* (*-11.6*, *-6.06*) | 5.50 (4.15, 7.31) | 540 (388, 742) | cost-saving[c] |
| WPR | 2.54 (2.20, 3.37) | *-2.99* (*-4.70*, *-1.58*) | 1.42 (1.04, 1.95) | 1830 (1250, 2640) | cost-saving[c] |

[a] Costs from the health system perspective include vaccination costs, tuberculosis testing and treatment costs, and antiretroviral treatment costs.
[b] Costs from the societal perspective include health system perspective costs, as well as patient non-medical costs and productivity losses.
[c] Both the point estimate and the interval estimates were cost-saving.
[d] High-TB, high-TB/HIV (HIV-associated TB), and high-MDR/RR-TB (multidrug/rifampicin-resistant TB) burden countries as defined by the World Health Organization [19].
[e] LIC: Gross national income (GNI) per capita of $1,085 or less; LMIC: GNI per capita of $1,086 to $4,225; UMIC: GNI per capita of $4,256 to $13,205 (World Bank 2021).

Note: All countries include 105 low- and middle-income countries analyzed. Values in parentheses represent equal-tailed 95% credible intervals. AFR = African region; AMR = Region of the Americas; EMR = Eastern Mediterranean region; EUR = European region; LIC = low-income; LMIC = lower middle-income; SEAR = Southeast Asian region; UMIC = upper middle-income; USD = United States dollar; WPR = Western Pacific region.

### S34. Discounted costs, disability-adjusted life-years (DALYs) averted, and cost-effectiveness of adolescent/adult tuberculosis vaccines: double-price scenario.

| Country grouping | Health system perspective[a] incremental cost (USD billions) | Societal perspective[b] incremental cost (USD billions) | DALYs averted (millions) | Health system cost (USD) per DALY averted | Societal cost (USD) per DALY averted |
|---|---|---|---|---|---|
| All countries | 58.8 (49.4, 77.7) | *-123* (*-142*, *-101*) | 94.8 (86.9, 103) | 622 (502, 821) | cost-saving[c] |
| High-TB burden[d] | 42.5 (35.5, 56.4) | *-125* (*-143*, *-106*) | 86.5 (78.5, 95.1) | 493 (395, 651) | cost-saving[c] |
| High-TB/HIV burden[d] | 26.3 (22.4, 33.5) | *-121* (*-137*, *-104*) | 77.1 (69.6, 85.3) | 343 (278, 443) | cost-saving[c] |
| High-MDR/RR-TB burden[d] | 39.4 (32.6, 52.9) | *-118* (*-135*, *-98.8*) | 76.9 (69.2, 85.3) | 513 (404, 693) | cost-saving[c] |
| **Income level[e]** | | | | | |
| LIC | 6.03 (5.28, 7.28) | *-0.27* (*-1.40*, 1.10) | 9.53 (8.40, 10.7) | 636 (523, 794) | cost-saving (cost-saving, 120) |
| LMIC | 28.6 (24.6, 35.9) | *-107* (*-121*, *-92.9*) | 78.2 (70.8, 86.3) | 368 (302, 464) | cost-saving[c] |
| UMIC | 24.1 (18.8, 36.1) | *-15.3* (*-25.1*, *-1.26*) | 7.15 (5.89, 8.83) | 3410 (2370, 5480) | cost-saving[c] |
| **World region** | | | | | |
| AFR | 11.3 (9.88, 13.6) | *-52.2* (*-62.0*, *-44.0*) | 38.9 (35.0, 42.8) | 290 (243, 364) | cost-saving[c] |
| AMR | 5.94 (4.70, 8.66) | 1.92 (0.56, 4.76) | 0.73 (0.65, 0.81) | 8190 (6260, 12000) | 2660 (747, 6430) |
| EMR | 6.60 (5.72, 8.14) | *-0.11* (*-1.92*, 1.83) | 6.61 (5.22, 8.24) | 1010 (761, 1350) | cost-saving (cost-saving, 332) |
| EUR | 3.31 (2.63, 4.80) | 0.59 (*-0.17*, 2.06) | 0.50 (0.45, 0.56) | 6580 (5060, 9860) | 1180 (cost-saving, 4280) |
| SEAR | 15.9 (13.5, 20.1) | *-64.8* (*-76.1*, *-53.3*) | 41.7 (35.8, 48.3) | 384 (300, 509) | cost-saving[c] |
| WPR | 15.8 (12.4, 23.4) | *-8.04* (*-12.7*, 0.01) | 6.37 (5.55, 7.31) | 2490 (1850, 3710) | cost-saving (cost-saving, 1.43) |

[a] Costs from the health system perspective include vaccination costs, tuberculosis testing and treatment costs, and antiretroviral treatment costs.
[b] Costs from the societal perspective include health system perspective costs, as well as patient non-medical costs and productivity losses.
[c] Both the point estimate and the interval estimates were cost-saving.
[d] High-TB, high-TB/HIV (HIV-associated TB), and high-MDR/RR-TB (multidrug/rifampicin-resistant TB) burden countries as defined by the World Health Organization [19].
[e] LIC: Gross national income (GNI) per capita of $1,085 or less; LMIC: GNI per capita of $1,086 to $4,225; UMIC: GNI per capita of $4,256 to $13,205 (World Bank 2021).
Note: All countries include 105 low- and middle-income countries analyzed. Values in parentheses represent equal-tailed 95% credible intervals. AFR = African region; AMR = Region of the Americas; EMR = Eastern Mediterranean region; EUR = European region; LIC = low-income; LMIC = lower middle-income; SEAR = Southeast Asian region; UMIC = upper middle-income; USD = United States dollar; WPR = Western Pacific region.

### S35. Discounted costs, disability-adjusted life-years (DALYs) averted, and cost-effectiveness of infant tuberculosis vaccines: high-middle-tier-price scenario.

| Country grouping | Health system perspective[a] incremental cost (USD billions) | Societal perspective[b] incremental cost (USD billions) | DALYs averted (millions) | Health system cost (USD) per DALY averted | Societal cost (USD) per DALY averted |
|---|---|---|---|---|---|
| All countries | 11.3 (9.86, 14.3) | *-23.8* (*-29.0*, *-18.7*) | 18.0 (15.6, 20.8) | 630 (516, 823) | cost-saving[c] |
| High-TB burden[d] | 7.38 (6.48, 9.37) | *-25.8* (*-30.8*, *-21.0*) | 16.6 (14.2, 19.5) | 446 (362, 579) | cost-saving[c] |
| High-TB/HIV burden[d] | 4.33 (3.74, 5.59) | *-24.2* (*-29.2*, *-19.8*) | 14.5 (12.2, 17.3) | 300 (237, 402) | cost-saving[c] |
| High-MDR/RR-TB burden[d] | 6.82 (6.01, 8.63) | *-24.2* (*-29.1*, *-19.4*) | 14.7 (12.4, 17.4) | 468 (372, 610) | cost-saving[c] |
| **Income level[e]** | | | | | |
| LIC | 1.12 (0.96, 1.44) | *-0.19* (*-0.48*, 0.15) | 2.02 (1.70, 2.46) | 558 (431, 737) | cost-saving (cost-saving, 77.2) |
| LMIC | 4.59 (3.95, 5.96) | *-22.4* (*-27.1*, *-18.1*) | 14.9 (12.6, 17.6) | 311 (245, 420) | cost-saving[c] |
| UMIC | 5.56 (4.92, 7.09) | *-1.19* (*-3.27*, 0.72) | 1.12 (0.85, 1.50) | 5090 (3570, 7190) | cost-saving (cost-saving, 832) |
| **World region** | | | | | |
| AFR | 2.50 (2.15, 3.23) | *-13.3* (*-17.0*, *-10.3*) | 9.34 (7.72, 11.3) | 271 (205, 364) | cost-saving[c] |
| AMR | 1.35 (1.18, 1.73) | 0.95 (0.76, 1.33) | 0.06 (0.05, 0.08) | 21400 (16700, 27900) | 15100 (10400, 21500) |
| EMR | 1.57 (1.38, 1.94) | *-0.10* (*-0.81*, 0.50) | 1.64 (1.15, 2.25) | 984 (666, 1450) | cost-saving (cost-saving, 422) |
| EUR | 0.75 (0.62, 1.04) | 0.52 (0.39, 0.81) | 0.04 (0.03, 0.04) | 19400 (15200, 27600) | 13500 (9320, 21700) |
| SEAR | 1.63 (1.35, 2.21) | *-9.86* (*-12.9*, *-7.34*) | 5.50 (4.15, 7.31) | 303 (209, 458) | cost-saving[c] |
| WPR | 3.48 (3.14, 4.31) | *-2.05* (*-3.76*, *-0.64*) | 1.42 (1.04, 1.95) | 2510 (1750, 3510) | cost-saving[c] |

[a] Costs from the health system perspective include vaccination costs, tuberculosis testing and treatment costs, and antiretroviral treatment costs.
[b] Costs from the societal perspective include health system perspective costs, as well as patient non-medical costs and productivity losses.
[c] Both the point estimate and the interval estimates were cost-saving.
[d] High-TB, high-TB/HIV (HIV-associated TB), and high-MDR/RR-TB (multidrug/rifampicin-resistant TB) burden countries as defined by the World Health Organization [19].
[e] LIC: Gross national income (GNI) per capita of $1,085 or less; LMIC: GNI per capita of $1,086 to $4,225; UMIC: GNI per capita of $4,256 to $13,205 (World Bank 2021).
Note: All countries include 105 low- and middle-income countries analyzed. Values in parentheses represent equal-tailed 95% credible intervals. AFR = African region; AMR = Region of the Americas; EMR = Eastern Mediterranean region; EUR = European region; LIC = low-income; LMIC = lower middle-income; SEAR = Southeast Asian region; UMIC = upper middle-income; USD = United States dollar; WPR = Western Pacific region.

**S36. Discounted costs, disability-adjusted life-years (DALYs) averted, and cost-effectiveness of adolescent/adult tuberculosis vaccines: high-middle-tier-price scenario.**

| Country grouping | Health system perspective[a] incremental cost (USD billions) | Societal perspective[b] incremental cost (USD billions) | DALYs averted (millions) | Health system cost (USD) per DALY averted | Societal cost (USD) per DALY averted |
|---|---|---|---|---|---|
| All countries | 54.7 (45.2, 73.5) | *-127* (*-146*, *-105*) | 94.8 (86.9, 103) | 578 (461, 778) | cost-saving[c] |
| High-TB burden[d] | 38.4 (31.4, 52.3) | *-129* (*-147*, *-110*) | 86.5 (78.5, 95.1) | 445 (351, 603) | cost-saving[c] |
| High-TB/HIV burden[d] | 19.1 (15.1, 26.2) | *-128* (*-144*, *-112*) | 77.1 (69.6, 85.3) | 248 (189, 345) | cost-saving[c] |
| High-MDR/RR-TB burden[d] | 36.6 (29.8, 50.1) | *-121* (*-138*, *-102*) | 76.9 (69.2, 85.3) | 477 (371, 655) | cost-saving[c] |
| **Income level[e]** | | | | | |
| LIC | 3.73 (2.98, 4.98) | *-2.57* (*-3.70*, *-1.20*) | 9.53 (8.40, 10.7) | 394 (301, 535) | cost-saving[c] |
| LMIC | 19.6 (15.5, 26.8) | *-116* (*-131*, *-102*) | 78.2 (70.8, 86.3) | 251 (191, 348) | cost-saving[c] |
| UMIC | 31.4 (26.0, 43.3) | *-8.11* (*-17.9*, 5.96) | 7.15 (5.89, 8.83) | 4440 (3240, 6570) | cost-saving (cost-saving, 884) |
| **World region** | | | | | |
| AFR | 7.23 (5.85, 9.59) | *-56.2* (*-66.1*, *-48.0*) | 38.9 (35.0, 42.8) | 186 (145, 255) | cost-saving[c] |
| AMR | 6.98 (5.73, 9.70) | 2.96 (1.60, 5.80) | 0.73 (0.65, 0.81) | 9630 (7590, 13500) | 4100 (2110, 7860) |
| EMR | 5.92 (5.04, 7.47) | *-0.79* (*-2.59*, 1.16) | 6.61 (5.22, 8.24) | 908 (674, 1230) | cost-saving (cost-saving, 210) |
| EUR | 3.74 (3.06, 5.23) | 1.02 (0.26, 2.49) | 0.50 (0.45, 0.56) | 7440 (5900, 10700) | 2040 (488, 5240) |
| SEAR | 9.65 (7.28, 13.9) | *-71.1* (*-82.4*, *-59.6*) | 41.7 (35.8, 48.3) | 233 (164, 344) | cost-saving[c] |
| WPR | 21.1 (17.7, 28.7) | *-2.72* (*-7.39*, 5.33) | 6.37 (5.55, 7.31) | 3330 (2600, 4560) | cost-saving (cost-saving, 837) |

[a] Costs from the health system perspective include vaccination costs, tuberculosis testing and treatment costs, and antiretroviral treatment costs.
[b] Costs from the societal perspective include health system perspective costs, as well as patient non-medical costs and productivity losses.
[c] Both the point estimate and the interval estimates were cost-saving.
[d] High-TB, high-TB/HIV (HIV-associated TB), and high-MDR/RR-TB (multidrug/rifampicin-resistant TB) burden countries as defined by the World Health Organization [19].
[e] LIC: Gross national income (GNI) per capita of $1,085 or less; LMIC: GNI per capita of $1,086 to $4,225; UMIC: GNI per capita of $4,256 to $13,205 (World Bank 2021).
Note: All countries include 105 low- and middle-income countries analyzed. Values in parentheses represent equal-tailed 95% credible intervals. AFR = African region; AMR = Region of the Americas; EMR = Eastern Mediterranean region; EUR = European region; LIC = low-income; LMIC = lower middle-income; SEAR = Southeast Asian region; UMIC = upper middle-income; USD = United States dollar; WPR = Western Pacific region.

**S37. Percentage of population that live in countries where vaccination was cost-effective from the health system perspective compared to percentage of gross domestic product per capita thresholds, comparing alternative vaccine price scenarios.**

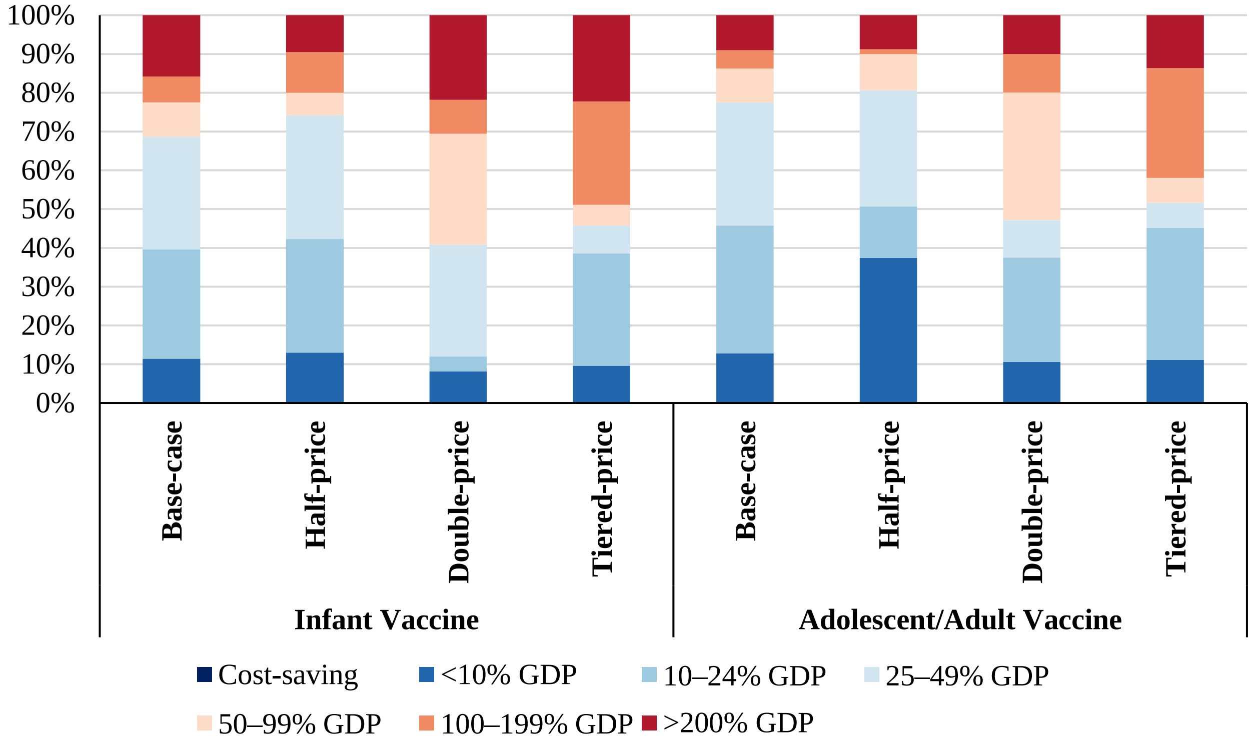


Note: Countries include 105 low- and middle-income countries analyzed. Population includes vaccinated individuals 2028–2050. Gross domestic product per capita estimates from 2020. GDP = gross domestic product per capita.

**S38. Percentage of countries where vaccination was cost-effective from the health system perspective compared to percentage of gross domestic product per capita thresholds, comparing alternative vaccine price scenarios.**

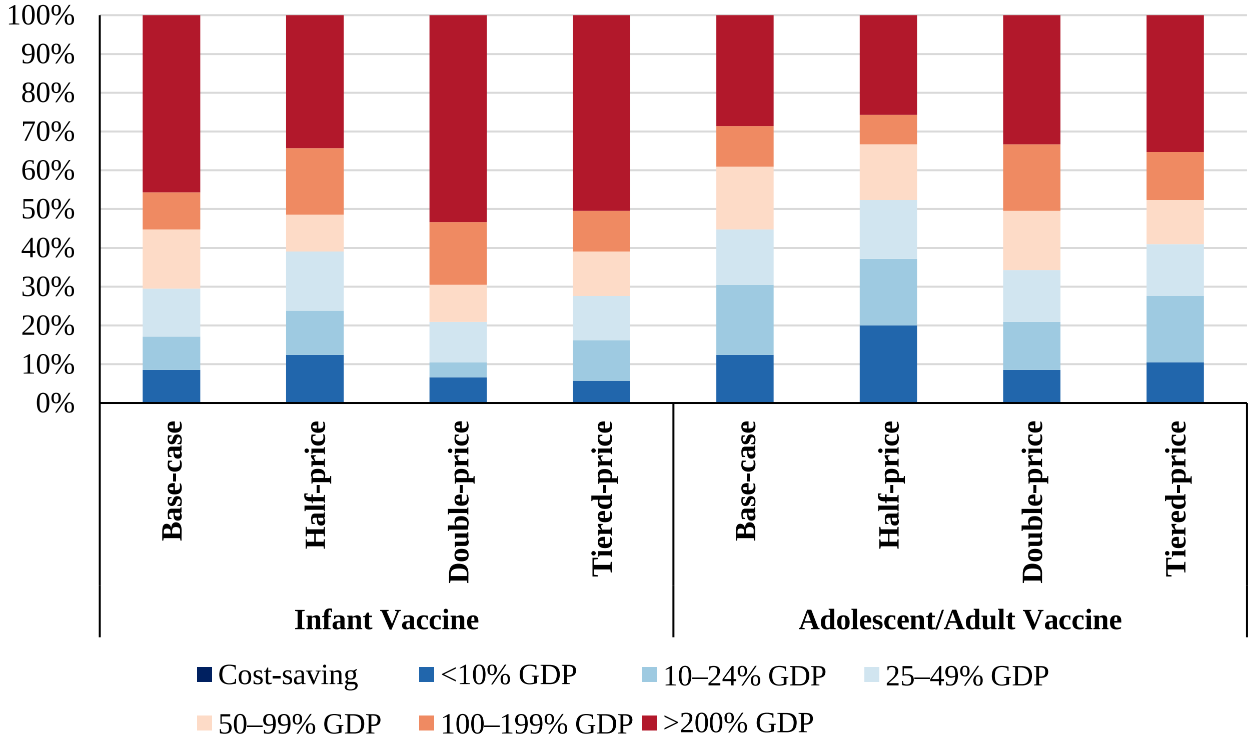


Note: Countries include 105 low- and middle-income countries analyzed. Gross domestic product per capita estimates from 2020. GDP = gross domestic product per capita.

### S39. Discounted costs, disability-adjusted life-years (DALYs) averted, and cost-effectiveness of infant tuberculosis vaccines: two-dose scenario.

| Country grouping | Health system perspective[a] incremental cost (USD billions) | Societal perspective[b] incremental cost (USD billions) | DALYs averted (millions) | Health system cost (USD) per DALY averted | Societal cost (USD) per DALY averted |
|---|---|---|---|---|---|
| All countries | 15.5 (12.7, 21.4) | *-19.6* (*-25.2*, *-12.7*) | 18.0 (15.6, 20.8) | 865 (671, 1220) | cost-saving[c] |
| High-TB burden[d] | 10.5 (8.73, 14.5) | *-22.7* (*-28.1*, *-16.9*) | 16.6 (14.2, 19.5) | 635 (492, 896) | cost-saving[c] |
| High-TB/HIV burden[d] | 7.41 (6.23, 9.94) | *-21.1* (*-26.3*, *-16.5*) | 14.5 (12.2, 17.3) | 513 (399, 714) | cost-saving[c] |
| High-MDR/RR-TB burden[d] | 9.26 (7.66, 13.0) | *-21.8* (*-27.0*, *-16.3*) | 14.7 (12.4, 17.4) | 635 (484, 916) | cost-saving[c] |
| **Income level[e]** | | | | | |
| LIC | 2.23 (1.92, 2.87) | 0.92 (0.53, 1.54) | 2.02 (1.70, 2.46) | 1110 (859, 1470) | 464 (227, 815) |
| LMIC | 8.09 (6.80, 10.8) | *-18.9* (*-23.8*, *-14.2*) | 14.9 (12.6, 17.6) | 548 (428, 763) | cost-saving[c] |
| UMIC | 5.18 (3.90, 8.23) | *-1.57* (*-4.02*, 1.70) | 1.12 (0.85, 1.50) | 4740 (2970, 7730) | cost-saving (cost-saving, 1680) |
| **World region** | | | | | |
| AFR | 4.84 (4.14, 6.31) | *-10.9* (*-14.6*, *-7.71*) | 9.34 (7.72, 11.3) | 524 (397, 711) | cost-saving[c] |
| AMR | 1.36 (1.03, 2.13) | 0.96 (0.62, 1.73) | 0.06 (0.05, 0.08) | 21600 (15200, 33700) | 15300 (8920, 27500) |
| EMR | 2.13 (1.75, 2.87) | 0.46 (*-0.32*, 1.32) | 1.64 (1.15, 2.25) | 1340 (875, 2040) | 321 (cost-saving, 1020) |
| EUR | 0.86 (0.60, 1.44) | 0.63 (0.38, 1.21) | 0.04 (0.03, 0.04) | 22200 (15000, 37600) | 16300 (9150, 31600) |
| SEAR | 3.29 (2.74, 4.49) | *-8.21* (*-11.3*, *-5.51*) | 5.50 (4.15, 7.31) | 610 (423, 919) | cost-saving[c] |
| WPR | 3.02 (2.34, 4.68) | *-2.51* (*-4.35*, *-0.50*) | 1.42 (1.04, 1.95) | 2180 (1390, 3540) | cost-saving[c] |

[a] Costs from the health system perspective include vaccination costs, tuberculosis testing and treatment costs, and antiretroviral treatment costs.
[b] Costs from the societal perspective include health system perspective costs, as well as patient non-medical costs and productivity losses.
[c] Both the point estimate and the interval estimates were cost-saving.
[d] High-TB, high-TB/HIV (HIV-associated TB), and high-MDR/RR-TB (multidrug/rifampicin-resistant TB) burden countries as defined by the World Health Organization [19].
[e] LIC: Gross national income (GNI) per capita of $1,085 or less; LMIC: GNI per capita of $1,086 to $4,225; UMIC: GNI per capita of $4,256 to $13,205 (World Bank 2021).
Note: All countries include 105 low- and middle-income countries analyzed. Values in parentheses represent equal-tailed 95% credible intervals. AFR = African region; AMR = Region of the Americas; EMR = Eastern Mediterranean region; EUR = European region; LIC = low-income; LMIC = lower middle-income; SEAR = Southeast Asian region; UMIC = upper middle-income; USD = United States dollar; WPR = Western Pacific region.

**S40. Discounted costs, disability-adjusted life-years (DALYs) averted, and cost-effectiveness of adolescent/adult tuberculosis vaccines: two-dose scenario.**

| Country grouping | Health system perspective[a] incremental cost (USD billions) | Societal perspective[b] incremental cost (USD billions) | DALYs averted (millions) | Health system cost (USD) per DALY averted | Societal cost (USD) per DALY averted |
|---|---|---|---|---|---|
| All countries | 75.9 (57.0, 114) | -106 (-132, -66.9) | 94.8 (86.9, 103) | 802 (591, 1200) | cost-saving[c] |
| High-TB burden[d] | 55.1 (41.1, 82.8) | -112 (-134, -82.8) | 86.5 (78.5, 95.1) | 639 (463, 964) | cost-saving[c] |
| High-TB/HIV burden[d] | 34.0 (26.3, 48.4) | -113 (-131, -92.8) | 77.1 (69.6, 85.3) | 442 (328, 629) | cost-saving[c] |
| High-MDR/RR-TB burden[d] | 51.1 (37.7, 78.2) | -106 (-128, -77.1) | 76.9 (69.2, 85.3) | 667 (477, 1030) | cost-saving[c] |
| **Income level[e]** | | | | | |
| LIC | 7.56 (6.05, 10.1) | 1.25 (-0.49, 3.75) | 9.53 (8.40, 10.7) | 796 (613, 1080) | 134 (cost-saving, 406) |
| LMIC | 36.8 (28.8, 51.2) | -98.9 (-115, -80.9) | 78.2 (70.8, 86.3) | 472 (357, 671) | cost-saving[c] |
| UMIC | 31.6 (21.0, 55.6) | -7.88 (-22.0, 18.7) | 7.15 (5.89, 8.83) | 4470 (2710, 8370) | cost-saving (cost-saving, 2850) |
| **World region** | | | | | |
| AFR | 14.3 (11.6, 19.0) | -49.2 (-59.3, -39.9) | 38.9 (35.0, 42.8) | 368 (286, 502) | cost-saving[c] |
| AMR | 7.71 (5.22, 13.2) | 3.70 (1.16, 9.26) | 0.73 (0.65, 0.81) | 10600 (7060, 18100) | 5110 (1590, 12700) |
| EMR | 8.39 (6.67, 11.5) | 1.68 (-0.64, 5.13) | 6.61 (5.22, 8.24) | 1290 (914, 1860) | 270 (cost-saving, 826) |
| EUR | 4.34 (2.97, 7.32) | 1.62 (0.23, 4.50) | 0.50 (0.45, 0.56) | 8620 (5790, 14900) | 3220 (440, 9270) |
| SEAR | 20.5 (15.9, 29.1) | -60.2 (-72.3, -46.9) | 41.7 (35.8, 48.3) | 495 (359, 722) | cost-saving[c] |
| WPR | 20.7 (13.9, 35.8) | -3.13 (-10.6, 12.6) | 6.37 (5.55, 7.31) | 3260 (2130, 5710) | cost-saving (cost-saving, 2030) |

[a] Costs from the health system perspective include vaccination costs, tuberculosis testing and treatment costs, and antiretroviral treatment costs.
[b] Costs from the societal perspective include health system perspective costs, as well as patient non-medical costs and productivity losses.
[c] Both the point estimate and the interval estimates were cost-saving.
[d] High-TB, high-TB/HIV (HIV-associated TB), and high-MDR/RR-TB (multidrug/rifampicin-resistant TB) burden countries as defined by the World Health Organization [19].
[e] LIC: Gross national income (GNI) per capita of $1,085 or less; LMIC: GNI per capita of $1,086 to $4,225; UMIC: GNI per capita of $4,256 to $13,205 (World Bank 2021).
Note: All countries include 105 low- and middle-income countries analyzed. Values in parentheses represent equal-tailed 95% credible intervals. AFR = African region; AMR = Region of the Americas; EMR = Eastern Mediterranean region; EUR = European region; LIC = low-income; LMIC = lower middle-income; SEAR = Southeast Asian region; UMIC = upper middle-income; USD = United States dollar; WPR = Western Pacific region.

### S41. Discounted costs, disability-adjusted life-years (DALYs) averted, and cost-effectiveness of infant tuberculosis vaccines: EndTB baseline scenario.

| Country grouping | Health system perspective[a] incremental cost (USD billions) | Societal perspective[b] incremental cost (USD billions) | DALYs averted (millions) | Health system cost (USD) per DALY averted | Societal cost (USD) per DALY averted |
|---|---|---|---|---|---|
| All countries | 8.01 (6.63, 10.9) | 4.24 (0.98, 7.83) | 1.73 (0.93, 3.21) | 5150 (2290, 9370) | 2940 (297, 7010) |
| High-TB burden[d] | 5.47 (4.59, 7.48) | 1.91 (-*1.32*, 4.74) | 1.60 (0.80, 3.10) | 3890 (1620, 7200) | 1610 (cost-saving, 5010) |
| High-TB/HIV burden[d] | 3.88 (3.30, 5.14) | 0.86 (-*2.11*, 2.99) | 1.36 (0.61, 2.77) | 3320 (1340, 6540) | 1070 (cost-saving, 4250) |
| High-MDR/RR-TB burden[d] | 4.84 (4.03, 6.71) | 1.52 (-*1.61*, 4.22) | 1.39 (0.62, 2.92) | 4050 (1550, 8180) | 1600 (cost-saving, 5530) |
| **Income level[e]** | | | | | |
| LIC | 1.14 (0.99, 1.46) | 1.02 (0.81, 1.34) | 0.18 (0.08, 0.35) | 7420 (3140, 15200) | 6750 (2470, 14400) |
| LMIC | 4.24 (3.60, 5.64) | 1.84 (-*0.92*, 3.88) | 1.33 (0.58, 2.87) | 3760 (1400, 7570) | 1940 (cost-saving, 5740) |
| UMIC | 2.63 (1.99, 4.16) | 1.37 (-*0.45*, 3.21) | 0.22 (0.06, 0.51) | 16500 (4300, 48100) | 10600 (cost-saving, 41100) |
| **World region** | | | | | |
| AFR | 2.50 (2.15, 3.24) | 0.49 (-*2.16*, 2.00) | 0.98 (0.41, 2.39) | 3100 (985, 6220) | 1040 (cost-saving, 4270) |
| AMR | 0.69 (0.52, 1.07) | 0.65 (0.47, 1.04) | 0.006 (0.003, 0.012) | 125000 (53400, 292000) | 119000 (47100, 285000) |
| EMR | 1.10 (0.91, 1.48) | 0.92 (0.38, 1.34) | 0.17 (0.02, 0.61) | 12300 (1740, 45900) | 11200 (629, 44200) |
| EUR | 0.44 (0.31, 0.73) | 0.39 (0.24, 0.69) | 0.007 (0.003, 0.014) | 79600 (27400, 190000) | 73500 (20300, 184000) |
| SEAR | 1.74 (1.46, 2.33) | 0.85 (-*0.45*, 1.70) | 0.42 (0.11, 1.09) | 5770 (1460, 17500) | 3560 (cost-saving, 15200) |
| WPR | 1.55 (1.22, 2.39) | 0.93 (-*0.19*, 1.94) | 0.15 (0.03, 0.49) | 16100 (2940, 49500) | 11600 (cost-saving, 44800) |

[a] Costs from the health system perspective include vaccination costs, tuberculosis testing and treatment costs, and antiretroviral treatment costs.
[b] Costs from the societal perspective include health system perspective costs, as well as patient non-medical costs and productivity losses.
[c] Both the point estimate and the interval estimates were cost-saving.
[d] High-TB, high-TB/HIV (HIV-associated TB), and high-MDR/RR-TB (multidrug/rifampicin-resistant TB) burden countries as defined by the World Health Organization [19].
[e] LIC: Gross national income (GNI) per capita of $1,085 or less; LMIC: GNI per capita of $1,086 to $4,225; UMIC: GNI per capita of $4,256 to $13,205 (World Bank 2021).
Note: All countries include 105 low- and middle-income countries analyzed. Values in parentheses represent equal-tailed 95% credible intervals. AFR = African region; AMR = Region of the Americas; EMR = Eastern Mediterranean region; EUR = European region; LIC = low-income; LMIC = lower middle-income; SEAR = Southeast Asian region; UMIC = upper middle-income; USD = United States dollar; WPR = Western Pacific region.

**S42. Discounted costs, disability-adjusted life-years (DALYs) averted, and cost-effectiveness of adolescent/adult tuberculosis vaccines: EndTB baseline scenario.**

| Country grouping | Health system perspective[a] incremental cost (USD billions) | Societal perspective[b] incremental cost (USD billions) | DALYs averted (millions) | Health system cost (USD) per DALY averted | Societal cost (USD) per DALY averted |
|---|---|---|---|---|---|
| All countries | 39.3 (29.8, 58.3) | 0.86 (-16.3, 23.3) | 18.7 (14.0, 25.8) | 2152 (1361, 3414) | 90.3 (cost-saving, 1340) |
| High-TB burden[d] | 28.7 (21.6, 42.6) | -5.96 (-21.6, 11.0) | 16.8 (12.2, 23.9) | 1756 (1075, 2879) | cost-saving (cost-saving, 737) |
| High-TB/HIV burden[d] | 17.9 (14.0, 25.2) | -11.8 (-25.3, -0.7) | 14.6 (10.3, 21.4) | 1269 (757, 1978) | cost-saving[c] |
| High-MDR/RR-TB burden[d] | 26.6 (20, 40.2) | -5.99 (-21.5, 11.1) | 15.0 (10.6, 21.9) | 1842 (1085, 3073) | cost-saving (cost-saving, 843) |
| **Income level[e]** | | | | | |
| LIC | 3.87 (3.12, 5.13) | 2.70 (1.75, 3.97) | 1.76 (1.23, 2.66) | 2289 (1388, 3458) | 1630 (686, 2770) |
| LMIC | 19.4 (15.4, 26.7) | -4.30 (-16.2, 5.78) | 14.4 (10.0, 21.5) | 1404 (840, 2139) | cost-saving (cost-saving, 486) |
| UMIC | 16.0 (10.7, 28.0) | 2.45 (-7.80, 15.7) | 2.58 (1.75, 4.10) | 6501 (3118, 12245) | 1240 (cost-saving, 7030) |
| **World region** | | | | | |
| AFR | 7.40 (6.07, 9.76) | -6.82 (-16.9, -0.89) | 7.54 (5.25, 11.7) | 1028 (580, 1562) | cost-saving[c] |
| AMR | 3.90 (2.66, 6.63) | 2.71 (1.45, 5.54) | 0.23 (0.19, 0.28) | 17186 (11143, 30784) | 12000 (5960, 25500) |
| EMR | 4.32 (3.43, 5.92) | 2.89 (0.77, 4.75) | 1.37 (0.74, 3.07) | 3578 (1294, 6409) | 2530 (256, 5360) |
| EUR | 2.18 (1.51, 3.67) | 1.07 (0.10, 2.58) | 0.20 (0.15, 0.28) | 11292 (6457, 20462) | 5700 (406, 14700) |
| SEAR | 10.9 (8.56, 15.2) | -3.33 (-13.2, 3.89) | 7.73 (4.83, 13.3) | 1513 (745, 2562) | cost-saving (cost-saving, 717) |
| WPR | 10.6 (7.12, 18.1) | 4.34 (-1.19, 12.7) | 1.63 (1.08, 2.87) | 6896 (3108, 13146) | 3030 (cost-saving, 9240) |

[a] Costs from the health system perspective include vaccination costs, tuberculosis testing and treatment costs, and antiretroviral treatment costs.

[b] Costs from the societal perspective include health system perspective costs, as well as patient non-medical costs and productivity losses.

[c] Both the point estimate and the interval estimates were cost-saving.

[d] High-TB, high-TB/HIV (HIV-associated TB), and high-MDR/RR-TB (multidrug/rifampicin-resistant TB) burden countries as defined by the World Health Organization [19].

[e] LIC: Gross national income (GNI) per capita of $1,085 or less; LMIC: GNI per capita of $1,086 to $4,225; UMIC: GNI per capita of $4,256 to $13,205 (World Bank 2021).

Note: All countries include 105 low- and middle-income countries analyzed. Values in parentheses represent equal-tailed 95% credible intervals. AFR = African region; AMR = Region of the Americas; EMR = Eastern Mediterranean region; EUR = European region; LIC = low-income; LMIC = lower middle-income; SEAR = Southeast Asian region; UMIC = upper middle-income; USD = United States dollar; WPR = Western Pacific region.

**S43. Discounted costs, disability-adjusted life-years (DALYs) averted, and cost-effectiveness of infant tuberculosis vaccines: lifelong duration of protection scenario.**

| Country grouping | Health system perspective[a] incremental cost (USD billions) | Societal perspective[b] incremental cost (USD billions) | DALYs averted (millions) | Health system cost (USD) per DALY averted | Societal cost (USD) per DALY averted |
|---|---|---|---|---|---|
| All countries | 7.40 (5.96, 10.4) | *-51.3* (*-59.4*, *-43.6*) | 30.0 (26.1, 34.5) | 248 (191, 356) | cost-saving[c] |
| High-TB burden[d] | 4.90 (3.99, 6.86) | *-50.7* (*-58.7*, *-43.1*) | 27.7 (23.8, 32.3) | 178 (136, 254) | cost-saving[c] |
| High-TB/HIV burden[d] | 3.42 (2.80, 4.68) | *-44.4* (*-52.5*, *-37.2*) | 24.3 (20.5, 28.7) | 142 (109, 200) | cost-saving[c] |
| High-MDR/RR-TB burden[d] | 4.29 (3.47, 6.07) | *-47.7* (*-55.5*, *-40.1*) | 24.5 (20.7, 28.9) | 177 (134, 261) | cost-saving[c] |
| **Income level[e]** | | | | | |
| LIC | 1.10 (0.95, 1.42) | *-1.06* (*-1.47*, *-0.66*) | 3.33 (2.80, 4.00) | 333 (258, 441) | cost-saving[c] |
| LMIC | 3.74 (3.07, 5.07) | *-41.1* (*-48.6*, *-34.4*) | 24.7 (21, 29.1) | 152 (117, 215) | cost-saving[c] |
| UMIC | 2.56 (1.92, 4.07) | *-9.17* (*-12.7*, *-6.42*) | 1.95 (1.48, 2.63) | 1340 (835, 2200) | cost-saving[c] |
| **World region** | | | | | |
| AFR | 2.33 (1.97, 3.05) | *-24.2* (*-30.1*, *-19.3*) | 15.6 (12.9, 18.6) | 151 (114, 207) | cost-saving[c] |
| AMR | 0.68 (0.52, 1.07) | *-0.003* (*-0.23*, 0.38) | 0.11 (0.09, 0.13) | 6270 (4400, 9770) | 19.2 (cost-saving, 3620) |
| EMR | 1.02 (0.82, 1.38) | *-1.73* (*-2.77*, *-0.85*) | 2.70 (1.90, 3.70) | 388 (249, 599) | cost-saving[c] |
| EUR | 0.43 (0.30, 0.72) | 0.03 (*-0.12*, 0.32) | 0.07 (0.06, 0.08) | 6480 (4370, 11000) | 473 (cost-saving, 4830) |
| SEAR | 1.49 (1.20, 2.08) | *-17.5* (*-22.5*, *-13.5*) | 9.11 (6.89, 12.0) | 167 (113, 257) | cost-saving[c] |
| WPR | 1.45 (1.11, 2.27) | *-7.89* (*-10.6*, *-5.75*) | 2.40 (1.76, 3.26) | 621 (396, 1020) | cost-saving[c] |

[a] Costs from the health system perspective include vaccination costs, tuberculosis testing and treatment costs, and antiretroviral treatment costs.

[b] Costs from the societal perspective include health system perspective costs, as well as patient non-medical costs and productivity losses.

[c] Both the point estimate and the interval estimates were cost-saving.

[d] High-TB, high-TB/HIV (HIV-associated TB), and high-MDR/RR-TB (multidrug/rifampicin-resistant TB) burden countries as defined by the World Health Organization [19].

[e] LIC: Gross national income (GNI) per capita of $1,085 or less; LMIC: GNI per capita of $1,086 to $4,225; UMIC: GNI per capita of $4,256 to $13,205 (World Bank 2021).

Note: All countries include 105 low- and middle-income countries analyzed. Values in parentheses represent equal-tailed 95% credible intervals. AFR = African region; AMR = Region of the Americas; EMR = Eastern Mediterranean region; EUR = European region; LIC = low-income; LMIC = lower middle-income; SEAR = Southeast Asian region; UMIC = upper middle-income; USD = United States dollar; WPR = Western Pacific region.

**S44. Discounted costs, disability-adjusted life-years (DALYs) averted, and cost-effectiveness of adolescent/adult tuberculosis vaccines: lifelong duration of protection scenario.**

| Country grouping | Health system perspective[a] incremental cost (USD billions) | Societal perspective[b] incremental cost (USD billions) | DALYs averted (millions) | Health system cost (USD) per DALY averted | Societal cost (USD) per DALY averted |
|---|---|---|---|---|---|
| All countries | 34.0 (24.5, 53.1) | *-232* (*-256*, *-205*) | 138 (127, 150) | 246 (173, 382) | cost-saving[c] |
| High-TB burden[d] | 24.0 (16.9, 38.0) | *-221* (*-244*, *-197*) | 126 (116, 138) | 190 (131, 298) | cost-saving[c] |
| High-TB/HIV burden[d] | 14.3 (10.1, 21.4) | *-200* (*-222*, *-179*) | 113 (102, 124) | 127 (87.7, 192) | cost-saving[c] |
| High-MDR/RR-TB burden[d] | 21.9 (15.1, 35.4) | *-208* (*-231*, *-184*) | 112 (102, 123) | 196 (131, 315) | cost-saving[c] |
| **Income level[e]** | | | | | |
| LIC | 3.65 (2.89, 4.90) | *-5.47* (*-6.84*, *-3.97*) | 13.8 (12.1, 15.4) | 266 (203, 363) | cost-saving[c] |
| LMIC | 15.5 (11.3, 22.8) | *-180* (*-200*, *-161*) | 113 (103, 125) | 137 (96.3, 204) | cost-saving[c] |
| UMIC | 14.8 (9.48, 26.8) | *-46.5* (*-60.5*, *-30.3*) | 11.2 (9.23, 13.9) | 1340 (795, 2580) | cost-saving[c] |
| **World region** | | | | | |
| AFR | 6.53 (5.10, 8.80) | *-87.3* (*-102*, *-75.3*) | 57.0 (51.5, 62.5) | 115 (88.0, 160) | cost-saving[c] |
| AMR | 3.78 (2.53, 6.50) | *-2.41* (*-3.90*, 0.49) | 1.13 (1.01, 1.26) | 3360 (2220, 5790) | cost-saving (cost-saving, 404) |
| EMR | 3.88 (3.00, 5.44) | *-5.98* (*-8.47*, *-3.59*) | 9.71 (7.69, 12.0) | 405 (280, 599) | cost-saving[c] |
| EUR | 1.94 (1.25, 3.41) | *-2.29* (*-3.15*, *-0.76*) | 0.77 (0.69, 0.86) | 2530 (1610, 4570) | cost-saving[c] |
| SEAR | 8.42 (5.92, 12.7) | *-107* (*-123*, *-91.2*) | 60.2 (52.0, 69.2) | 141 (92.1, 214) | cost-saving[c] |
| WPR | 9.47 (5.97, 17.0) | *-26.6* (*-32.0*, *-18.4*) | 9.62 (8.39, 11.1) | 988 (610, 1810) | cost-saving[c] |

[a] Costs from the health system perspective include vaccination costs, tuberculosis testing and treatment costs, and antiretroviral treatment costs.
[b] Costs from the societal perspective include health system perspective costs, as well as patient non-medical costs and productivity losses.
[c] Both the point estimate and the interval estimates were cost-saving.
[d] High-TB, high-TB/HIV (HIV-associated TB), and high-MDR/RR-TB (multidrug/rifampicin-resistant TB) burden countries as defined by the World Health Organization [19].
[e] LIC: Gross national income (GNI) per capita of $1,085 or less; LMIC: GNI per capita of $1,086 to $4,225; UMIC: GNI per capita of $4,256 to $13,205 (World Bank 2021).
Note: All countries include 105 low- and middle-income countries analyzed. Values in parentheses represent equal-tailed 95% credible intervals. AFR = African region; AMR = Region of the Americas; EMR = Eastern Mediterranean region; EUR = European region; LIC = low-income; LMIC = lower middle-income; SEAR = Southeast Asian region; UMIC = upper middle-income; USD = United States dollar; WPR = Western Pacific region.

### S45. Discounted costs, disability-adjusted life-years (DALYs) averted, and cost-effectiveness of adolescent/adult tuberculosis vaccines: 75% efficacy of adolescent/adult vaccine scenario.

| Country grouping | Health system perspective[a] incremental cost (USD billions) | Societal perspective[b] incremental cost (USD billions) | DALYs averted (millions) | Health system cost (USD) per DALY averted | Societal cost (USD) per DALY averted |
|---|---|---|---|---|---|
| All countries | 34.3 (24.9, 53.4) | *-230* (*-255*, *-203*) | 138 (127, 150) | 250 (175, 387) | cost-saving[c] |
| High-TB burden[d] | 24.3 (17.1, 38.3) | *-219* (*-243*, *-195*) | 125 (114, 138) | 194 (134, 302) | cost-saving[c] |
| High-TB/HIV burden[d] | 14.4 (10.3, 21.6) | *-199* (*-221*, *-177*) | 112 (101, 123) | 130 (89.8, 194) | cost-saving[c] |
| High-MDR/RR-TB burden[d] | 22.2 (15.4, 35.6) | *-206* (*-229*, *-182*) | 112 (101, 123) | 200 (134, 319) | cost-saving[c] |
| **Income level[e]** | | | | | |
| LIC | 3.65 (2.90, 4.92) | *-5.46* (*-6.83*, *-3.97*) | 13.8 (12.2, 15.4) | 266 (203, 362) | cost-saving[c] |
| LMIC | 15.7 (11.5, 23.0) | *-180* (*-201*, *-162*) | 113 (103, 125) | 139 (97.9, 206) | cost-saving[c] |
| UMIC | 15.0 (9.64, 26.9) | *-43.7* (*-57.0*, *-27.7*) | 10.7 (8.80, 13.2) | 1420 (855, 2720) | cost-saving[c] |
| **World region** | | | | | |
| AFR | 6.61 (5.19, 8.87) | *-86.1* (*-100*, *-74.2*) | 56.5 (51.1, 62.0) | 117 (89.8, 163) | cost-saving[c] |
| AMR | 3.80 (2.55, 6.51) | *-2.13* (*-3.59*, 0.76) | 1.08 (0.96, 1.20) | 3540 (2330, 6060) | cost-saving (cost-saving, 670) |
| EMR | 3.91 (3.03, 5.46) | *-5.76* (*-8.15*, *-3.44*) | 9.52 (7.52, 11.8) | 416 (290, 615) | cost-saving[c] |
| EUR | 1.96 (1.28, 3.44) | *-2.08* (*-2.93*, *-0.54*) | 0.75 (0.67, 0.83) | 2640 (1690, 4750) | cost-saving[c] |
| SEAR | 8.48 (5.98, 12.7) | *-108* (*-124*, *-92.0*) | 60.5 (52.3, 69.8) | 141 (92.3, 215) | cost-saving[c] |
| WPR | 9.58 (6.10, 17.1) | *-25.4* (*-30.8*, *-17.4*) | 9.36 (8.15, 10.7) | 1030 (642, 1870) | cost-saving[c] |

[a] Costs from the health system perspective include vaccination costs, tuberculosis testing and treatment costs, and antiretroviral treatment costs.

[b] Costs from the societal perspective include health system perspective costs, as well as patient non-medical costs and productivity losses.

[c] Both the point estimate and the interval estimates were cost-saving.

[d] High-TB, high-TB/HIV (HIV-associated TB), and high-MDR/RR-TB (multidrug/rifampicin-resistant TB) burden countries as defined by the World Health Organization [19].

[e] LIC: Gross national income (GNI) per capita of $1,085 or less; LMIC: GNI per capita of $1,086 to $4,225; UMIC: GNI per capita of $4,256 to $13,205 (World Bank 2021).

Note: All countries include 105 low- and middle-income countries analyzed. Values in parentheses represent equal-tailed 95% credible intervals. AFR = African region; AMR = Region of the Americas; EMR = Eastern Mediterranean region; EUR = European region; LIC = low-income; LMIC = lower middle-income; SEAR = Southeast Asian region; UMIC = upper middle-income; USD = United States dollar; WPR = Western Pacific region.

### References


1. Clark RA, Mukandavire C, Portnoy A, Weerasuriya CK, Deol A, Scarponi D, et al. The impact of alternative delivery strategies for novel tuberculosis vaccines in low- and middle-income countries: a modelling study. medRxiv; 2022. doi: https://doi.org/10.1101/2022.04.16.22273762.
2. Frascella B, Richards AS, Sossen B, Emery JC, Odone A, Law I, et al. Subclinical Tuberculosis Disease-A Review and Analysis of Prevalence Surveys to Inform Definitions, Burden, Associations, and Screening Methodology. Clin Infect Dis. 2021;73(3):e830-e41.
3. Emery JC, Richards AS, Dale KD, McQuaid CF, White RG, Denholm JT, et al. Self-clearance of Mycobacterium tuberculosis infection: implications for lifetime risk and population at-risk of tuberculosis disease. Proc Biol Sci. 2021;288(1943):20201635.
4. World Health Organization. Guidelines for treatment of drug-susceptible tuberculosis and patient care: 2017 update. 2017. World Health Organization. https://apps.who.int/iris/handle/10665/255052. License: CC BY-NC-SA 3.0 IGO.
5. World Health Organization. Treatment of drug-susceptible tuberculosis: rapid communication. Geneva: World Health Organization. 2021. https://apps.who.int/iris/handle/10665/341729 (accessed Jan 7, 2022). .
6. Kwan CK, Ernst JD. HIV and tuberculosis: a deadly human syndemic. Clin Microbiol Rev. 2011;24(2):351-76.
7. Vynnycky E, Fine PE. The natural history of tuberculosis: the implications of age-dependent risks of disease and the role of reinfection. Epidemiol Infect. 1997;119(2):183-201.
8. Suthar AB, Lawn SD, del Amo J, Getahun H, Dye C, Sculier D, et al. Antiretroviral therapy for prevention of tuberculosis in adults with HIV: a systematic review and meta-analysis. PLoS Med. 2012;9(7):e1001270.
9. Iskauskas A. hmer: History Matching and Emulation Package. R package. 17 May 2022. Available at: https://CRAN.R-project.org/package=hmer (accessed 12 July 2022).
10. Scarponi D, Iskauskas A, Clark RA, Vernon I, McKinley TJ, Goldstein M, et al. Demonstrating Multi-Country Calibration of a Tuberculosis Model Using New History Matching and Emulation Package - Hmer. medRxiv; 2022. doi: https://doi.org/10.1101/2022.05.13.22275052.
11. Jabot F, Faure T, Dumoulin N, Albert C. EasyABC: Efficient Approximate Bayesian Computation Sampling Schemes. R package. 2015. Available at: https://CRAN.R-project.org/package=EasyABC (accessed April 2022).
12. World Health Organization. WHO Preferred Product Characteristics for New Tuberculosis Vaccines. Geneva: World Health Organization. Licence: CC BY-NC-SA 3.0 IGO. 13 July 2018. https://www.who.int/publications/i/item/WHO-IVB-18.06 (accessed 18 October 2021).
13. Abubakar I, Pimpin L, Ariti C, Beynon R, Mangtani P, Sterne JA, et al. Systematic review and meta-analysis of the current evidence on the duration of protection by bacillus Calmette-Guérin vaccination against tuberculosis. Health Technol Assess. 2013;17(37):1-372, v-vi.
14. Blackwood JC, Cummings DA, Broutin H, Iamsirithaworn S, Rohani P. Deciphering the impacts of vaccination and immunity on pertussis epidemiology in Thailand. Proc Natl Acad Sci U S A. 2013;110(23):9595-600.

15. Lewnard JA, Grad YH. Vaccine waning and mumps re-emergence in the United States. Sci Transl Med. 2018;10(433).
16. Gavi The Vaccine Alliance. Country hub. Geneva: Gavi, The Vaccine Alliance. https://www.gavi.org/programmes-impact/country-hub (accessed 15 April 2022).
17. UNICEF. Vaccination and Immunization Statistics. https://data.unicef.org/topic/child-health/immunization/ (accessed March 7, 2022). .
18. Harris RC, Sumner T, Knight GM, Zhang H, White RG. Potential impact of tuberculosis vaccines in China, South Africa, and India. Sci Transl Med. 2020;12(564).
19. World Health Organization. WHO releases new global lists of high-burden countries for TB, HIV-associated TB and drug-resistant TB. 17 June 2021. https://www.who.int/news/item/17-06-2021-who-releases-new-global-lists-of-high-burden-countries-for-tb-hiv-associated-tb-and-drug-resistant-tb (accessed 1 July 2021).
20. Global Burden of Disease Collaborative Network. Global Burden of Disease Study 2019 (GBD 2019) Disability Weights. Seattle, United States of America: Institute for Health Metrics and Evaluation (IHME), 2020. doi: https://doi.org/10.6069/1W19-VX76 (accessed 23 August 2021).
21. Husereau D, Drummond M, Augustovski F, de Bekker-Grob E, Briggs AH, Carswell C, et al. Consolidated Health Economic Evaluation Reporting Standards 2022 (CHEERS 2022) Statement: Updated Reporting Guidance for Health Economic Evaluations. Clin Ther. 2022;44(2):158-68.
